# Inferring the sensitivity of wastewater metagenomic sequencing for early detection of viruses: a statistical modelling study

**DOI:** 10.1101/2023.12.22.23300450

**Authors:** Simon L. Grimm, Jeff T. Kaufman, Daniel P. Rice, Charles Whittaker, William J. Bradshaw, Michael R. McLaren

## Abstract

**Background:** Metagenomic sequencing of wastewater (W-MGS) can in principle detect any known or novel pathogen in a population. We quantify the sensitivity and cost of W-MGS for viral pathogen detection by jointly analysing W-MGS and epidemiological data for a range of human-infecting viruses.

**Methods:** Sequencing data from four studies were analysed to estimate the relative abundance (RA) of 11 human-infecting viruses. Corresponding prevalence and incidence estimates were obtained or calculated from academic and public-health reports. These estimates were combined using a hierarchical Bayesian model to predict RA at set prevalence or incidence values, allowing comparison across studies and viruses. These predictions were then used to estimate the sequencing depth and concomitant cost required for pathogen detection using W-MGS with or without use of a hybridization-capture enrichment panel.

**Findings:** After controlling for variation in local infection rates, relative abundance varied by orders of magnitude across studies for a given virus. For instance, a local SARS-CoV-2 weekly incidence of 1% corresponds to predicted SARS-CoV-2 relative abundance ranging from 3.8 × 10^−10^ to 2.4 × 10^−7^ across studies, translating to orders-of-magnitude variation in the cost of operating a system able to detect a SARS-CoV-2-like pathogen at a given sensitivity. Use of a respiratory virus enrichment panel in two studies dramatically increased predicted relative abundance of SARS-CoV-2, lowering yearly costs by 24-to 29-fold for a system able to detect a SARS-CoV-2-like pathogen before reaching 0.01% cumulative incidence.

**Interpretation:** The large variation in viral relative abundance after controlling for epidemiological factors indicates that other sources of inter-study variation, such as differences in sewershed hydrology and lab protocols, have a substantial impact on the sensitivity and cost of W-MGS. Well-chosen hybridization capture panels can dramatically increase sensitivity and reduce cost for viruses in the panel, but may reduce sensitivity to unknown or unexpected pathogens.

**Funding:** Wellcome Trust; Open Philanthropy; Musk Foundation

**Research In Context:** *Evidence before this study:* Numerous other studies have performed wastewater metagenomic sequencing (W-MGS), with a range of objectives. However, few have explicitly examined the performance of W-MGS as a monitoring tool. We searched PubMed between database inception and September 2024, using the search terms “MGS OR Metagenomic sequencing OR Metagenomics OR Shotgun sequencing” AND “Performance OR Precision OR Sensitivity OR Cost-effectiveness OR Effectiveness” AND “Virus OR Viral OR Virome” AND “Wastewater OR Sewage”. Among the 88 resulting studies, 17 focused specifically on viruses in wastewater. A 2023 UK study by Child and colleagues assessed hybridization-capture and untargeted sequencing of wastewater for genomic epidemiology, concluding that the former but not the latter provided sufficient coverage for effective variant tracking. However, they did find untargeted sequencing sufficient for presence/absence calls of human pathogens in wastewater, a finding supported by numerous other W-MGS studies. While several studies examined the effect of different W-MGS protocols on viral abundance and composition, none accounted for epidemiological or study effects, and none explicitly quantified the sensitivity and cost of W-MGS for viral detection.

*Added value of this study:* To our knowledge, this study provides the first quantitative assessment of the sensitivity and cost of untargeted and hybridization-capture W-MGS for pathogen surveillance. Linking a large corpus of public wastewater metagenomic sequencing with epidemiological data in a Bayesian model, we predict pathogen relative abundance in W-MGS data at set infection prevalence or incidence, and estimate concomitant read-depth and cost requirements for effective detection across different studies and viruses. Our flexible modelling framework provides a valuable tool for evaluation of sequencing-based surveillance in other contexts.

*Implications of all the available evidence:* The sensitivity of untargeted W-MGS varies greatly with pathogen and study design, and large gaps in our understanding remain for pathogens not present in our data. As untargeted W-MGS protocols undergo further improvements, our Bayesian modelling framework is an effective tool for evaluating the sensitivity of new protocols under different epidemiological conditions. While less pathogen-agnostic, hybridization capture can dramatically increase the sensitivity of W-MGS-based pathogen monitoring, and our findings support piloting it as a tool for biosurveillance of known viruses.

## Introduction

Many enteric and non-enteric viruses have been found to shed via human faeces into wastewater (1,2), and monitoring of viruses in wastewater has been used as a public health tool since the identification of poliovirus in sewage in 1939 (3). Today, wastewater surveillance is a mature technology deployed globally across high-, middle-, and low-income countries (4). Many such programs use qPCR to monitor the spread of pathogens like influenza A and SARS-CoV-2, while using amplicon sequencing to track viral variants (5).

However, these technologies can only detect a defined list of target pathogens. In contrast, untargeted wastewater metagenomic sequencing (W-MGS) and, to a lesser extent, hybridization-capture sequencing (e.g., VirCapSeq-VERT (6)) are able to detect a far wider range of viruses, including those that are unexpected or new to science.

However, despite this promise, W-MGS has so far seen little use in biosurveillance, due primarily to concerns about its sensitivity and cost (7,8). Several studies have qualitatively assessed W-MGS for viral pathogen detection (7,9) and genomic epidemiology (10). However, none have controlled for variation in community infection rates or quantitatively evaluated the sensitivity of untargeted W-MGS. This stands in contrast to targeted wastewater surveillance, which has seen more quantitative study (11–13).

To address this gap, our paper assesses the performance of untargeted and hybridization-capture wastewater sequencing for virus surveillance. To do so, we developed a hierarchical Bayesian model that combines publicly available metagenomic datasets with epidemiological data for a range of known viral pathogens, allowing us to estimate the relative abundance of a given virus in wastewater at a given prevalence or incidence. We then integrated these estimates with a parameterized cost model to estimate the cost of pathogen detection with W-MGS.

## Methods

### Wastewater sequencing data and bioinformatic analysis

To identify appropriate sequencing data for analysis, we performed a literature search for studies which (i) generated large (>100M read pairs), untargeted shotgun W-MGS datasets from raw treatment plant influent (ii) used sample preparation methods well-suited for broad enrichment of viruses, and (iii) were performed in regions and time periods for which good public-health data were available.

We selected three RNA-sequencing studies which fit all of these criteria: Crits-Christoph et al. 2021 (7), Rothman et al. 2021 (9), and Spurbeck et al. 2023 (14). While we were unable to find any DNA-sequencing studies that fulfilled all three criteria, we were still interested in assessing the capability of DNA sequencing to detect human-infecting viruses. Therefore, we included the DNA sequencing study by Brinch et al. 2020 (15), which fulfils criteria (i) and (iii).

All four of these studies conducted composite sampling of municipal influent (the three RNA studies all used 24-hour composite samples, while Brinch used 12-hour composites) and sequenced processed samples with paired-end Illumina technology. The three RNA sequencing studies were conducted in the United States, sampling wastewater from California and Ohio between 2020 and 2022. Brinch sampled wastewater in Copenhagen, Denmark from 2015 to 2018.

For these studies, we obtained sequencing reads from the European Nucleotide Archive (16) and identified virus reads with a custom computational pipeline using Bowtie2 (17) and Kraken2 (18) (appendix 5 p 23). Relative abundance of each virus was calculated as the number of high-quality, non-duplicate reads assigned to that virus divided by the total number of sequencing reads.

In addition to untargeted W-MGS data, Crits-Christoph and Rothman also sequenced samples that had undergone hybridization-capture enrichment with the Illumina Respiratory Virus Panel (RVP). Data from these samples underwent the same bioinformatic analysis as the untargeted samples from the same studies.

### Epidemiological data collation

We sought location-specific public health indicators (incidence or prevalence) for various viruses that could explain the variation in W-MGS relative abundance seen for each virus.

To select viruses for which to collate and generate incidence and prevalence estimates, we performed an exploratory literature review. We included viruses if they met the following criteria: (i) recognized public health importance based on morbidity or prevalence, and (ii) availability of incidence or prevalence estimates, or related data such as testing rates or seroprevalence measurements (appendix 2 p 9). Viruses were categorised as acute-infecting or chronic-infecting based on their typical infection duration and shedding patterns. Though some chronic-infecting viruses have distinct acute phases upon initial infection (such as chickenpox caused by VZV), reliable incidence data for these infections was often unavailable. Thus, we estimated weekly incidence for acute-infecting viruses, and prevalence for chronic-infecting viruses. Detailed methodologies for each virus’ epidemiological estimate are provided in appendix 4 (p 22).

Among acute-infecting viruses, we included SARS-CoV-2, influenza A/B, and norovirus GI/GII; respiratory syncytial virus (RSV) was excluded due to its near-complete suppression during the RNA sequencing study timeframes, which coincided with COVID-19 mitigation measures (19). Among chronic-infecting viruses, we included Human Immunodeficiency Virus (HIV, only accounting for individuals without viral suppression (20)), Hepatitis C Virus (HCV), Epstein-Barr Virus (EBV), Herpes Simplex Virus 1 (HSV-1), Cytomegalovirus (CMV), and Human Papillomavirus (HPV). Details on how we arrived at incidence and prevalence estimates can be found in appendix 2 (p 9) and appendix 4 (p 22).

### Statistical model and inference

We sought to infer the relationship between the public health indicator and relative abundance for a given virus and study. To do so, we developed a generative hierarchical Bayesian model that describes how the number of reads for a virus observed in a W-MGS sample depend on its current public health indicator, total reads from that sample, and a conversion factor *B* between the public health indicator and the virus’ relative abundance. We infer *B* separately for each study and virus, allowing for variation in *B* across sampling locations within a study. We implemented the model in the Stan probabilistic programming language^15^, using PyStan (21) (v3.10.0) for model fitting and evaluation. See (appendix 3 page 19) for details on derivation, implementation, fitting, and evaluation.

To aid interpretation of the results, we define a metric, *RA*(*x*%), equal to the expected relative abundance of a pathogen in W-MGS data when the corresponding public health indicator equals *x*%. For small values of *x*%, *RA*(*x*%)*≈B·x*%. All results are presented at 1% infection prevalence or weekly infection incidence totalling 1% of the population, denoted as RA_p_(1%), and RA_i_(1%), respectively. Figure S6 shows location-specific RA_i_(1%) derived from the hierarchical model’s location-specific coefficients *B* for each location *l* (appendix 3 p 20); otherwise, we report RA(1%) for the study using the study’s global coefficient *B*. We report posterior medians with 90% credible intervals.

### Estimating the cost of pathogen detection with W-MGS

To estimate the cost of pathogen detection with W-MGS, we focus on acute-infecting viruses for which we have weekly incidence data. We first estimate the required weekly sequencing depth (number of total reads) required to detect a given virus at a given cumulative incidence (appendix 3 p 20), then estimate the weekly cost required to achieve this sequencing depth (appendix 6 p 24). We operationally define detection as occurring when a given cumulative number of reads from the pathogen (the *detection threshold*) are observed. We set hypothetical detection thresholds ranging from 10 - 1000 total reads.

To estimate required sequencing depth, we derive the relationship between the detection threshold, weekly sequencing depth, and cumulative incidence at detection under our model for an emerging pandemic that follows a given incidence-to-RA relationship (appendix 3 p 20). We then use the relationship we inferred for particular viruses and studies to estimate the required weekly depth for various detection thresholds and cumulative incidence targets.

To estimate the cost associated with carrying out weekly sequencing to the required depth, we calculated the sum of two components (appendix 6 p 24). The first was a fixed per-sample processing cost associated with library preparation and other associated laboratory methods. The second was a per-read sequencing cost that scaled with sequencing depth (appendix 6 p 25); to calculate this component, we obtained published estimates of per-unit (lane or flow cell) costs for different Illumina sequencing platforms; calculated the number of units required for each platform to reach the required sequencing depth; calculated the cost for each platform as the product of the per-unit cost and the number of units required; and then took the minimum cost across platforms. For both components, cost estimates were based on published pricing information from the Harvard Bauer Core Facility and Illumina.

### Statement of ethical approval

No ethical approval was required for this study.

## Results

Analysing viral relative abundance in untargeted W-MGS data from 481 samples across four studies, we found that the median relative abundance of viruses differed by orders of magnitude between studies, from 6.3×10^−4^ in Spurbeck to 2.0×10^−2^ in Rothman. The relative abundance of human-infecting viruses also varied widely, albeit less so than that of total viruses, from 2.5×10^−6^ in Spurbeck to 1.8×10^−5^ in Crits-Christoph (figure 1a).

**Figure 1:**
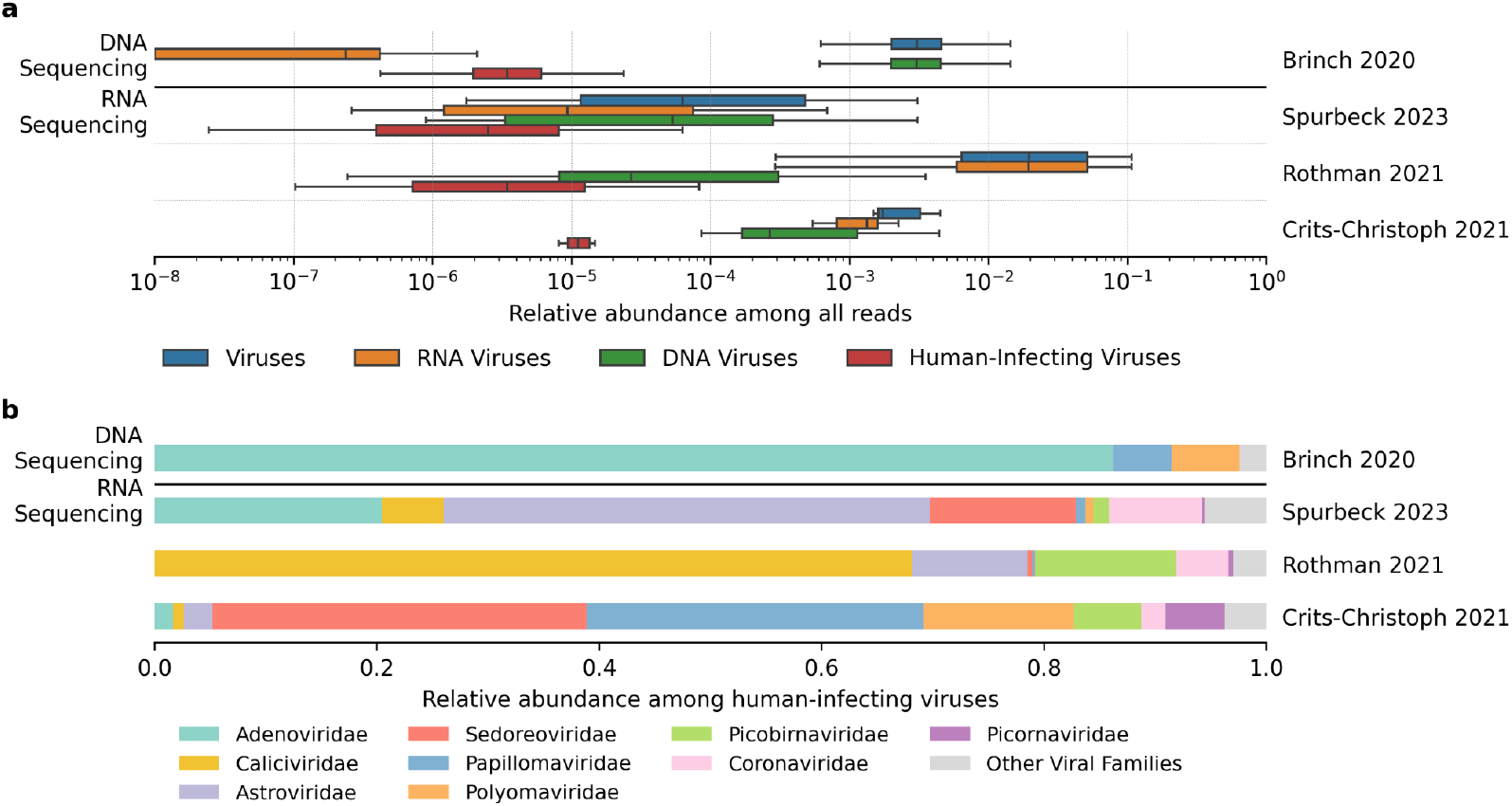
Viral abundance and taxonomic composition across wastewater metagenomics studies. **(a)** Relative abundance of all viruses (blue), RNA viruses (orange), human-infecting viruses (green), and DNA viruses (red), across samples for each included study. Values under 10^−8^ are equal to 0. **(b)** Taxonomic composition of human-infecting viruses in each study, displayed as the fraction of reads mapping to different virus families among all human-infecting virus reads.

The taxonomic composition of human-infecting viruses also varied dramatically between studies (figure 1b), even within the group of studies that performed RNA-sequencing. For example, the fraction of human-infecting virus reads mapping to Caliciviridae varied from 9.9 × 10^−3^ in Crits-Christoph to 6.8 × 10^−1^ in Rothman, while Sedoreoviridae varied from 4.4 × 10^−3^ in Rothman to 3.4 × 10^−1^ in Crits-Christoph.

Observed differences in viral abundance and composition could arise from a variety of factors, most notably the number of people infected with each virus contributing to the sewershed. To account for this, we collected infection prevalence and/or incidence estimates for 11 human-infecting viruses of recognised public-health importance, and used a hierarchical Bayesian model to generate normalised estimates of relative abundance for each study at a specific prevalence or incidence (Methods). Here we present the estimated relative abundance at 1% infection prevalence or weekly incidence in the study population (RAi(1%)); results for different epidemiological values vary proportionally.

We find large differences in estimated viral relative abundance between studies after controlling for these epidemiological indicators (appendix 1 p 3). The largest inter-study differences were observed for SARS-CoV-2 and norovirus, which showed inter-study variation in median RAi(1%) of roughly three orders of magnitude (figure 2a & appendix 1 p 4). These inter-study differences were not consistent in direction; for example, Rothman showed higher RAi(1%) than Crits-Christoph for norovirus GII, but lower for SARS-CoV-2. All three RNA-sequencing studies, however, were consistent in showing much higher median RAi(1%) for norovirus GI than SARS-CoV-2, consistent with the former’s status as a virus that spreads primarily via the faecal-oral route.

**Figure 2:**
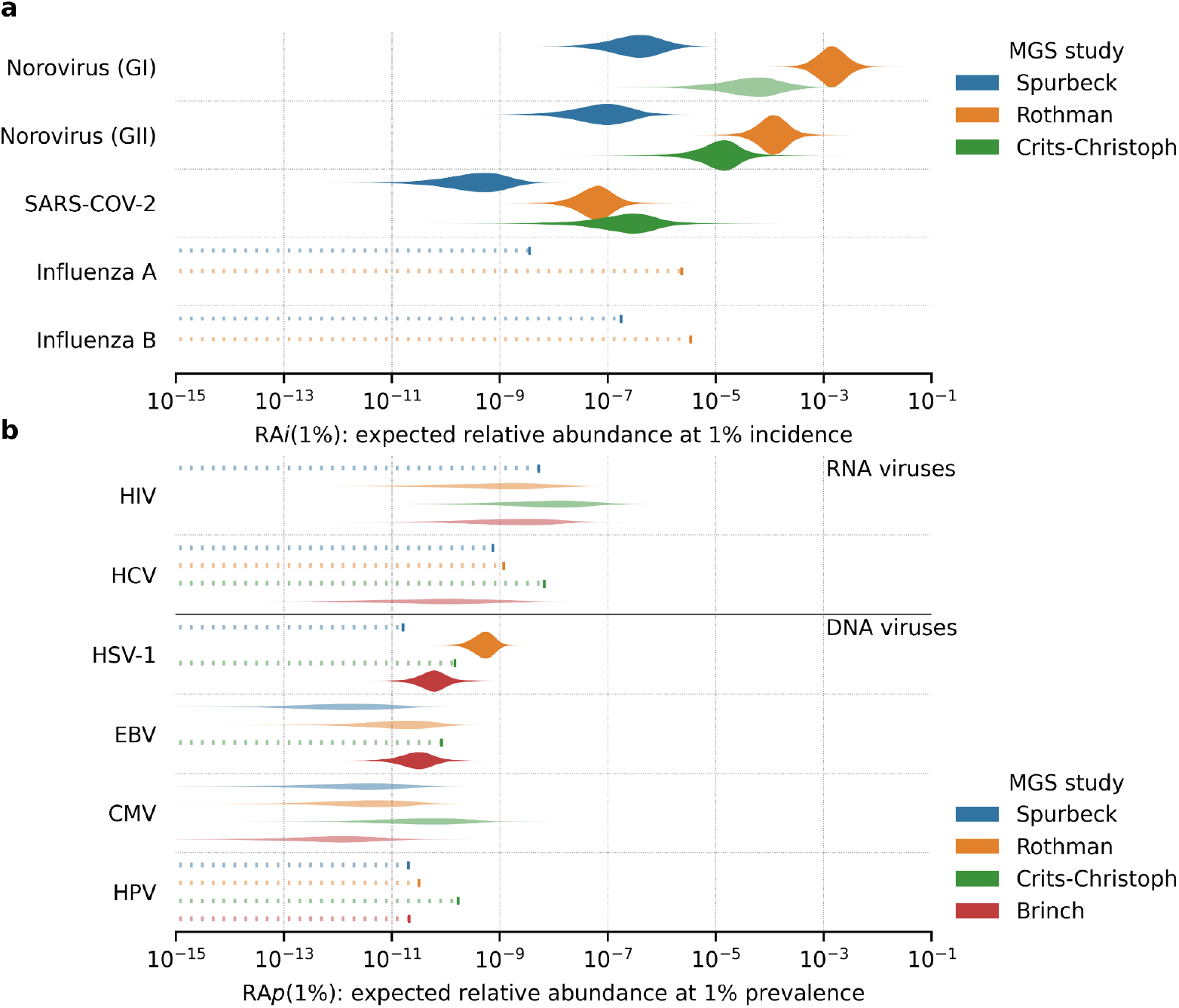
Study-level relative abundance (*RA*) estimates (untargeted W-MGS). **(a)** Predicted *RA* of acute viruses at 1% weekly incidence (*RA*_i_(1%)). Influenza RA_*i*_(1%) for Crits-Christoph is not displayed due to very low estimated incidence during the study period, rendering the results unreliable. No incidence estimates were created for Brinch (appendix 4 p 22). **(b)** Predicted *RA* of chronic viruses at 1% prevalence (*RA*_p_(1%)). Each violin represents the posterior predictive distribution of our Bayesian model for a specific study and virus. Transparent violins indicate predictions made with <10 reads mapping to the corresponding virus; dashed lines indicate predictions made with zero mapped reads, with the 95th percentile result indicated by a larger dash.

Influenza and several chronic viruses (e.g. HCV, HPV, HSV-1) showed very low abundance in wastewater data, with zero mapped reads in two to four studies. In these cases it was still possible to estimate an upper bound on RA(1%), representing the information we obtain from knowing that a study did *not* detect a virus at some sequencing depth. Such upper bounds differ between studies and viruses based on differences in total read count and epidemiological indicators, with higher incidence or prevalence resulting in lower upper-bound estimates of RA(1%).

For other chronic viruses that did appear consistently in the data, inter-study differences in RA(1%) were typically smaller than for acute RNA viruses, though still substantial; for example, the median RA(1%) of HIV (untreated) and EBV varied by roughly one order of magnitude across studies (figure 2b & appendix 1 p 4). In the case of HIV in particular, the measured RA values may be overestimates, as we were unable to rule out the presence of lentiviral cloning vectors among HIV-assigned reads.

In addition to the 481 untargeted W-MGS samples analysed above, two of the RNA studies, Rothman and Crits-Christoph, also included samples (266 and 11, respectively) that had undergone hybridization capture enrichment of a subset of human-infecting viruses with the Illumina Respiratory Virus Panel (RVP) (22). By repeating the above analysis on these samples, we were able to estimate the effect of this enrichment on viral relative abundance.

The Illumina RVP includes probes for roughly 40 human-infecting respiratory viruses, of which 3 (SARS-CoV-2, influenza A, influenza B) were included in the group of 11 viruses for which we collected public-health data.

SARS-CoV-2 was observed in a substantially larger fraction of hybrid-capture-enriched than unenriched samples in both studies (79% vs 24% in Rothman, 100% vs 71% in Crits-Christoph). On average, unadjusted relative abundance of SARS-CoV-2 increased 3-fold in Rothman and 80,000-fold in Crits-Christoph. Accounting for epidemiological indicators, median estimates of RA_i_(1%) for SARS-CoV-2 were 4.1 × 10^−6^ (1.1 × 10^−6^ to 1.7 × 10^−5^) and 1.7 × 10^−1^ (2.1 × 10^−2^ to 5.8 × 10^−1^) in hybridization-capture samples of Rothman and Crits-Christoph, a 65- and 750,000-fold increase over non-enriched samples (figure 3a, appendix 2 p 12). The reason for the dramatic difference in the apparent effectiveness of hybridization capture between studies, for both raw and adjusted relative-abundance estimates, is unclear.

**Figure 3:**
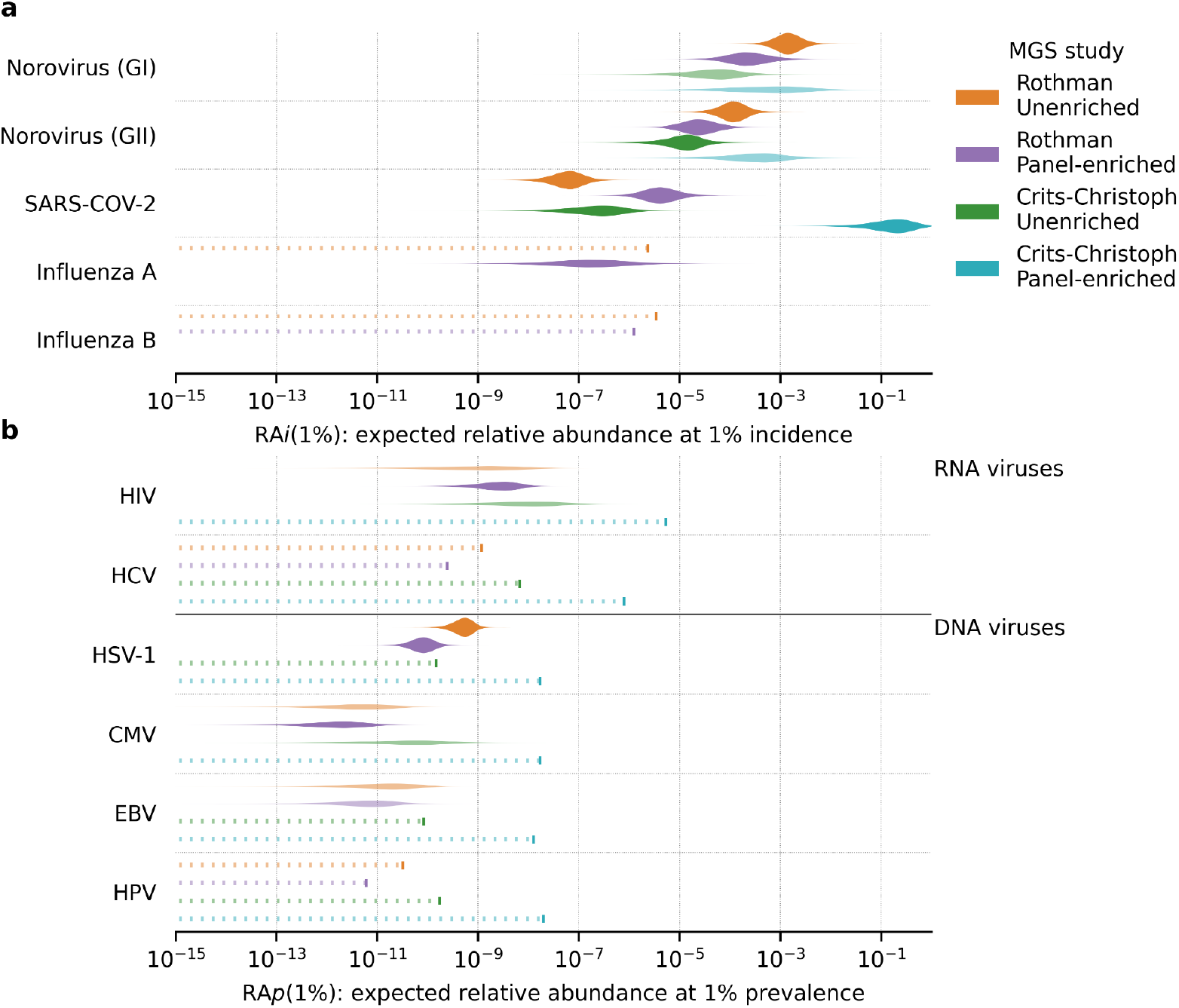
Study-level relative abundance (*RA*) estimates for Rothman and Crits-Christoph, unenriched and hybridization-capture-enriched samples. Influenza RAi(1%) for Crits-Christoph is not displayed (appendix 4, p 25). **(a)** Predicted RA of acute viruses at 1% weekly incidence (RAi(1%)). (b) Predicted *RA* of chronic viruses at 1% prevalence (*RA*_p_(1%)). Transparent violins indicate predictions made with <10 reads mapping to the corresponding virus; dashed lines indicate predictions made with zero mapped reads, with the 95th percentile result indicated by a larger dash.

Previously absent in all studies, we identified influenza A in 13 out of 266 hybrid-capture-enriched samples in Rothman, with a mean relative abundance of 7.4 × 10^−7^. Accounting for influenza incidence at the time of sampling, the median estimate of RA_i_(1%) for influenza A in Rothman was 1.6 × 10^−7^(7.9 × 10^−9^ - 2.4 × 10^−6^). This relatively low estimate involves high uncertainty due to the small number of influenza A reads (39) and low average weekly incidence in California during sampling (19 cases per 100,000 per week) (figure 3, appendix 1 p 3).

Among viruses not represented in the panel, no uniform pattern of increase or decrease in sample positivity or median relative abundance was observed. When accounting for differences in incidence and prevalence, median Norovirus GII RA_i_(1%) fell 5-fold in Rothman, but increased 25-fold in Crits-Christoph. For untreated HIV, RA_i_(1%) rose 2.5-fold in the enriched fraction of Rothman but was undetected in enriched Crits-Christoph samples, providing only an upper-bound estimate of 5.2 × 10^−6^ (appendix 2 p 12).

We next developed a simple statistical model to convert our estimates of the relationship between viral relative abundance and disease incidence into estimates of the required sequencing depth to detect a given virus at a given cumulative incidence. We considered a virus to be detected when the cumulative reads associated with that virus crossed a predefined threshold. Given this detection threshold we estimated the weekly sequencing depth *n* (appendix 3, p 21, figure 4).

**Figure 4:**
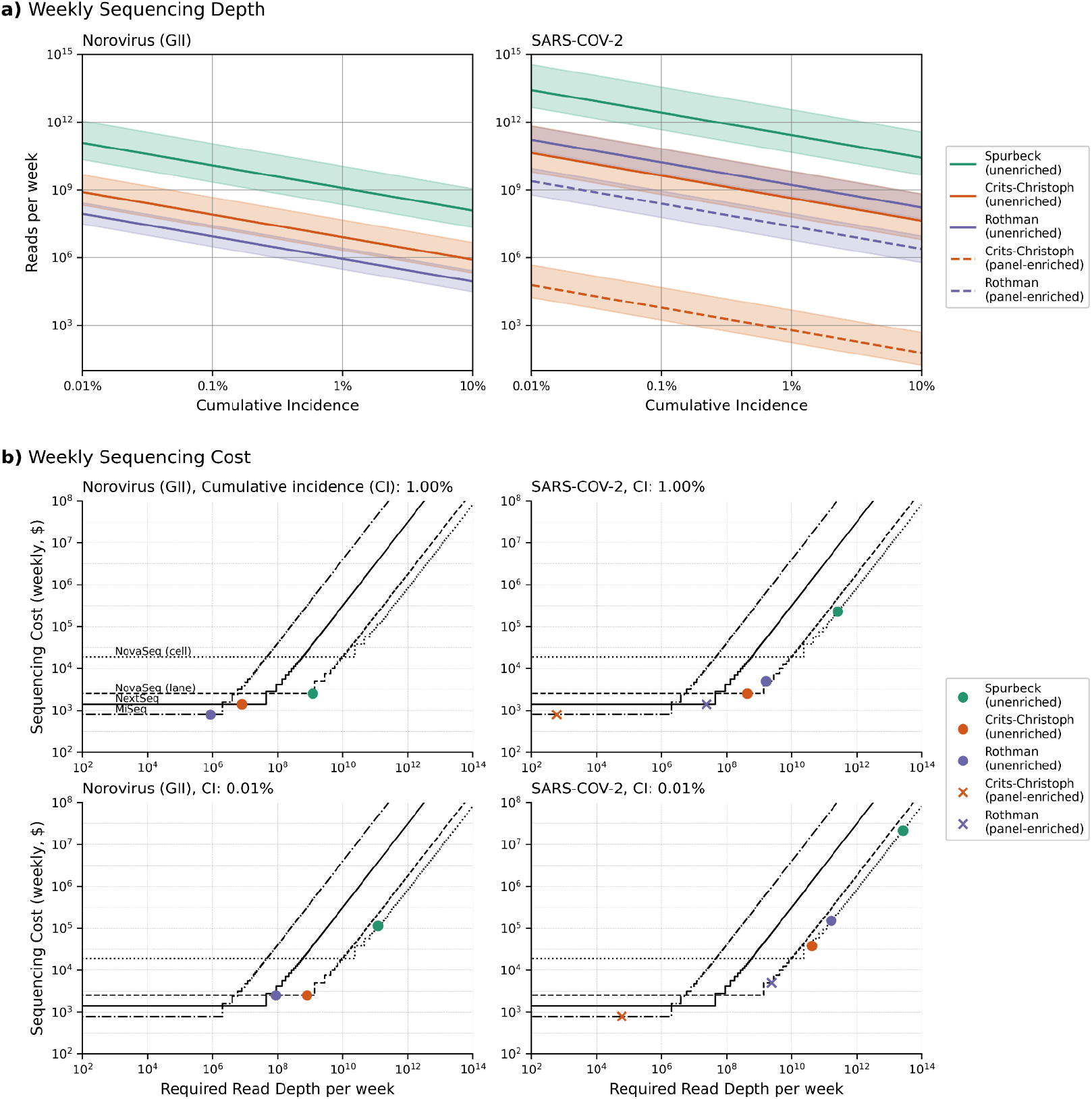
Weekly sequencing depth and cost. **a)** Predicted weekly reads required for Norovirus and SARS-CoV-2 detection at a given target cumulative incidence, given RAi(1%) median and 90% CI estimates for different studies and enrichment methods. **b)** Per-sample sequencing costs for detection of SARS-CoV-2 and Norovirus (GII) at different cumulative incidence levels, using different detection thresholds. Step functions show costs and sequencing depth for MiSeq (dash-dot line), NextSeq (solid line), NovaSeq flow cell lane (dashed line), and NovaSeq full flow cell (dotted line). Dots represent the required read depth and down-stream costs for different studies (Crits-Christoph, Rothman, Spurbeck) and sample types (enriched [circles], unenriched [crosses]). Sequencing depth modeling assumes a detection threshold of 100 reads.

The number of reads required varies substantially between different viruses and studies. For untargeted sequencing, we find that detection at 1% cumulative incidence, with a read threshold of 100 reads, requires 8.7 × 10^5^, 8.0 × 10^6^, and 1.2 × 10^9^ sequencing reads for norovirus GII and 1.7 × 10^9^, 4.3 × 10^8^, and 2.6 × 10^11^ for SARS-CoV-2 in Rothman, Crits-Christoph, and Spurbeck, respectively. Detection of SARS-CoV-2 via hybridization-capture sequencing requires fewer sequencing reads, with 2.4 × 10^7^ and 6.0 × 10^2^ reads in Rothman and Crits-Christoph respectively. Changing the detection threshold alters the required read depth proportionally.

Finally, we used our estimates of required sequencing depth to estimate the cost of W-MGS-based pathogen detection, combining estimates of per-sample and per-read costs for each dataset (Methods, appendix 1, pp 6, appendix 2, p 16, appendix 6, pp 24-25). We assume a detection threshold of 100 reads and a target cumulative incidence (TCI) of either 1% or 0.01%.

At 1% TCI, the predicted annual cost to detect a GII norovirus-like pathogen under Rothman, Crits-Christoph, and Spurbeck was $52,200, $86,800, and $141,000, respectively. Lowering the TCI to 0.01% increases these estimates to $141,000, $144,000, and $5.91 million (appendix 2 p 16, figure 4).

With the exception of the Spurbeck estimate at 0.01% TCI, these numbers are relatively affordable, reflecting the high abundance of faecal-oral viruses in municipal wastewater.

Estimates of detection costs for a SARS-CoV-2-like pathogen using untargeted W-MGS were both higher and more variable than for norovirus, primarily due to the low RA(1%) estimate for Spurbeck. At 1% TCI, the estimated cost under Rothman, Crits-Christoph, and Spurbeck was $271,000, $144,000, and $11.8 million. At 0.01% TCI, these costs increased to $7.87 million, $1.98 million, and over $1 billion respectively.

Unlike norovirus, SARS-CoV-2 is included in the Illumina RVP, and is consequently enriched in hybrid-capture-enriched samples from Rothman and Crits-Christoph. Using RA(1%) estimates derived from these samples, the estimated cost of detection of a SARS-CoV-2-like pathogen at 1% TCI falls to $101,000 for Rothman and only $69,100 for Crits-Christoph. At 0.01% TCI, the estimated cost increases to $287,000 for Rothman, but remains $69,100 for Crits-Christoph, requiring only a single MiSeq run per week at both TCIs. A well-chosen hybridization-capture strategy can thus dramatically reduce the predicted cost of W-MGS-based pathogen detection (appendix 1, pp 6, appendix 2, p 12).

## Discussion

Metagenomic surveillance of wastewater (W-MGS) could enable early detection of a wide range of known and unknown pathogens, strains, and variants without reliance on clinical presentation. However, any surveillance effort that hopes to detect novel viruses while they are still rare will need to sequence samples to high depth, incurring substantial sequencing costs. Estimating the scale of these costs is an important component of any attempt to evaluate W-MGS for biosurveillance.

We created a Bayesian model that integrates W-MGS data with epidemiological estimates to estimate expected relative abundance of different viral pathogens at a given incidence or prevalence. Applying this model to data from a range of studies, we find that predicted relative abundance varies widely across both pathogens and studies. Our results strongly suggest that non-epidemiological sources of inter-study variation have a substantial impact on the sensitivity of W-MGS. Important sources of variation include the properties of the sewershed itself: differences in the relative contribution of human waste versus other material (e.g. stormwater, agricultural waste, in-sewer microbial growth), transit time, and temperature can all affect microbial composition and viral decay. Choices made by the experimenter regarding sampling and processing methodologies (e.g. influent vs sludge, viral concentration method) are also likely to play a major role; more research into the specific effects of different protocol choices on W-MGS sensitivity will be an important part of its maturation as a biosurveillance tool.

The large variation in predicted relative abundance in W-MGS data across studies and pathogens led to correspondingly wide variation in the predicted cost of pathogen detection. For a target cumulative incidence (TCI) of 1% at a single site, predicted annual costs range from ~$50,000–140,000 for Norovirus to ~$140,000–12 million for SARS-CoV-2. Lowering the TCI to 0.01% increases the necessary costs to $140,000-2 million for Norovirus and >$2 million for SARS-CoV-2. However, enriching for SARS-CoV-2 via hybridization capture dramatically reduces the predicted cost at low TCI, with the annual cost falling from $2 million to less than $70,000 under Crits-Christoph and from $7 million to $290,000 under Rothman. Well-designed hybridization capture panels could thus significantly improve the feasibility of W-MGS-based biosurveillance.

While this study represents an important step forward in our understanding of W-MGS-based biosurveillance, key limitations remain. Most notably, there is the limited amount of available sequencing data from regions with robust public-health estimates. In total, the studies we selected for in-depth analysis comprised roughly 3B RNA-sequencing reads and 4B DNA-sequencing reads: a significant amount compared to most individual studies, but inadequate to robustly quantify viruses at very low relative abundances, such as influenza. Correspondingly, many viruses returned zero mapped reads in some or all of the included studies, a problem exacerbated by a lack of DNA studies that deliberately enriched for viruses. Future research should use significantly deeper sequencing data, generated with protocols optimized for viral W-MGS.

The available public health data also had important limitations. Many RNA viruses either had little epidemiological information available (metapneumovirus, rhinovirus) or near zero incidence during the periods covered by our data (respiratory syncytial virus), making robust estimates impractical. Where robust estimates were available, existing literature often presented data as point estimates without adequate or consistent representation of uncertainty. Converting data for different pathogens into common metrics required the use of a range of different conversion factors that also failed to adequately represent uncertainty. While our results align with prior expectations and enable meaningful comparisons across studies, improvements in available public-health data and estimation methods would substantially aid future research in this domain.

We see two major avenues for future research. First, our parameter estimates can be integrated into more advanced cost-effectiveness models that compare different pathogen detection approaches while incorporating other factors influencing W-MGS performance. Second, future work should improve parameter estimates by incorporating larger viral MGS datasets, higher-quality public-health data, and additional sequencing platforms. Generating and incorporating large, virally-enriched DNA-sequencing datasets from wastewater would be especially valuable. As sequencing costs continue to decline, such research will be central in determining under what circumstances untargeted W-MGS is suitable for routine deployment. In the meantime, hybrid-capture sequencing represents a lower-cost alternative that could substantially improve our ability to detect and mitigate future viral outbreaks.

## Data Availability

All data produced are available online at https://github.com/naobservatory/mgs-workflow/tree/2.1.0 (metagenomic data analysis pipeline) and https://github.com/naobservatory/p2ra-manuscript (epidemiological analysis, statistical models, figure generation). The output of our biocomputational pipeline is available here: https://doi.org/10.6084/m9.figshare.28395104.v1.

## Data sharing

All sequencing data in this study is available through the European Nucleotide Archive(16), using BioProject accessions listed in appendix 2 (p 7). The raw data used for estimating pathogen prevalence and incidence are listed in the respective Python script for each pathogen. All code used in this study can be found in two Github repositories, https://github.com/naobservatory/mgs-workflow/tree/2.1.0 (metagenomic data analysis pipeline) and https://github.com/naobservatory/p2ra-manuscript (epidemiological analysis, statistical models, figure generation). The output of our biocomputational pipeline is available here: https://doi.org/10.6084/m9.figshare.28395104.v1.

## Acknowledgements

S.L.G., J.T.K., D.P.R., W.J.B., and M.R.M. were funded for this research project by gifts from Open Philanthropy (to SecureBio) and the Musk Foundation (to MIT). S.L.G. was additionally supported through a grant by the Swiss Scholarship Foundation. C.W. was supported by Sir Henry Wellcome Postdoctoral Fellowship, reference 224190/Z/21/Z. We thank Asher Parker-Sartori for help obtaining public-health data and generating prevalence and incidence estimates. We also gratefully acknowledge Lenni Justen for providing helpful feedback on earlier drafts of this manuscript.

## Author contributions

J.T.K. and M.R.M. conceived the study; S.L.G. and J.T.K. performed literature reviews on pathogen incidence and prevalence and collected and evaluated epidemiological datasets with input from D.P.R.; J.T.K. and W.J.B. performed literature reviews on metagenomics data with feedback from M.R.M.; J.T.K. and W.J.B. imported and processed sequencing data; D.P.R. created the study’s statistical models with feedback from M.R.M., W.J.B., and J.T.K.; S.L.G., J.T.K., and D.P.R. created figures with feedback from W.J.B., M.R.M. and C.W.; S.L.G., W.J.B., M.R.M., J.T.K. & D.P.R. wrote the manuscript, with feedback from all authors.

