## Supplementary Information for "Inferring the sensitivity of wastewater metagenomic sequencing for early detection of viruses: a statistical modelling study"

### Table of Contents

**Table of Contents** **2**
Appendix 1: Supplementary figures 3
Figure S1: Prevalence and incidence of human-infecting viruses. 3
Figure S2: Inter-study variability in RA(1%). 4
Figure S3. Location-level relative abundance (RA1%) estimates for selected viruses. 5
Figure S4: Per-sample processing and sequencing cost. Sources for processing and sequencing costs are given in appendix 6, p 2. 6
Appendix 2: Supplementary tables 7
Table S1: Median relative abundance of all viruses and human-infecting viruses.
Table S2: Bioproject IDs for all included studies. 7
Table S3: Metadata for all included studies. 7
Table S4: Reasons for why no epidemiological estimates were created for Respiratory Syncytial Virus, Rhinovirus, Enteric Adenovirus, Metapneumovirus, and Hepatitis A Virus. 8
Table S5: Human-infecting viruses included in study. 9
Table S6: RA(1%) estimates for human-infecting viruses in unenriched sequencing data. 11
Table S7: Comparison of RA(1%) between hybridization-capture and unenriched samples. 13
Table S8: Weekly sequencing required for detection at 1% cumulative incidence and a detection threshold of 100 reads. 14
Table S9: Weekly sequencing required for detection at 1% cumulative incidence for hybridization capture samples. 15
Table S10: Estimated sequencing costs and depths for detection of SARS-CoV-2 and Norovirus (GII) at 1% and 0·01% cumulative incidence. 17
Table S11: Effect on varying pseudocounts (total positive cases in area over one week) on final RA(1%) values for acute-infecting pathogens. 17
Appendix 3: Statistical inference and required sequencing depth 17
Estimating the PH-to-RA conversion factor and RA(1%) 17
Incidence pseudocounts 19
Pathogens with zero reads 20
Model fitting and checking 20
Relationship between weekly sequencing depth and cumulative incidence at detection 20
Appendix 4: Epidemiological data 22
Epidemiological data 22
Virus selection 22
Data analysis 22
Appendix 5: Metagenomic data analysis 23
Appendix 6: Cost modeling 24
Overview 24
Per-sample processing costs 24
Per-sample sequencing costs 25
Supplementary references 26

#### Appendix 1: Supplementary figures

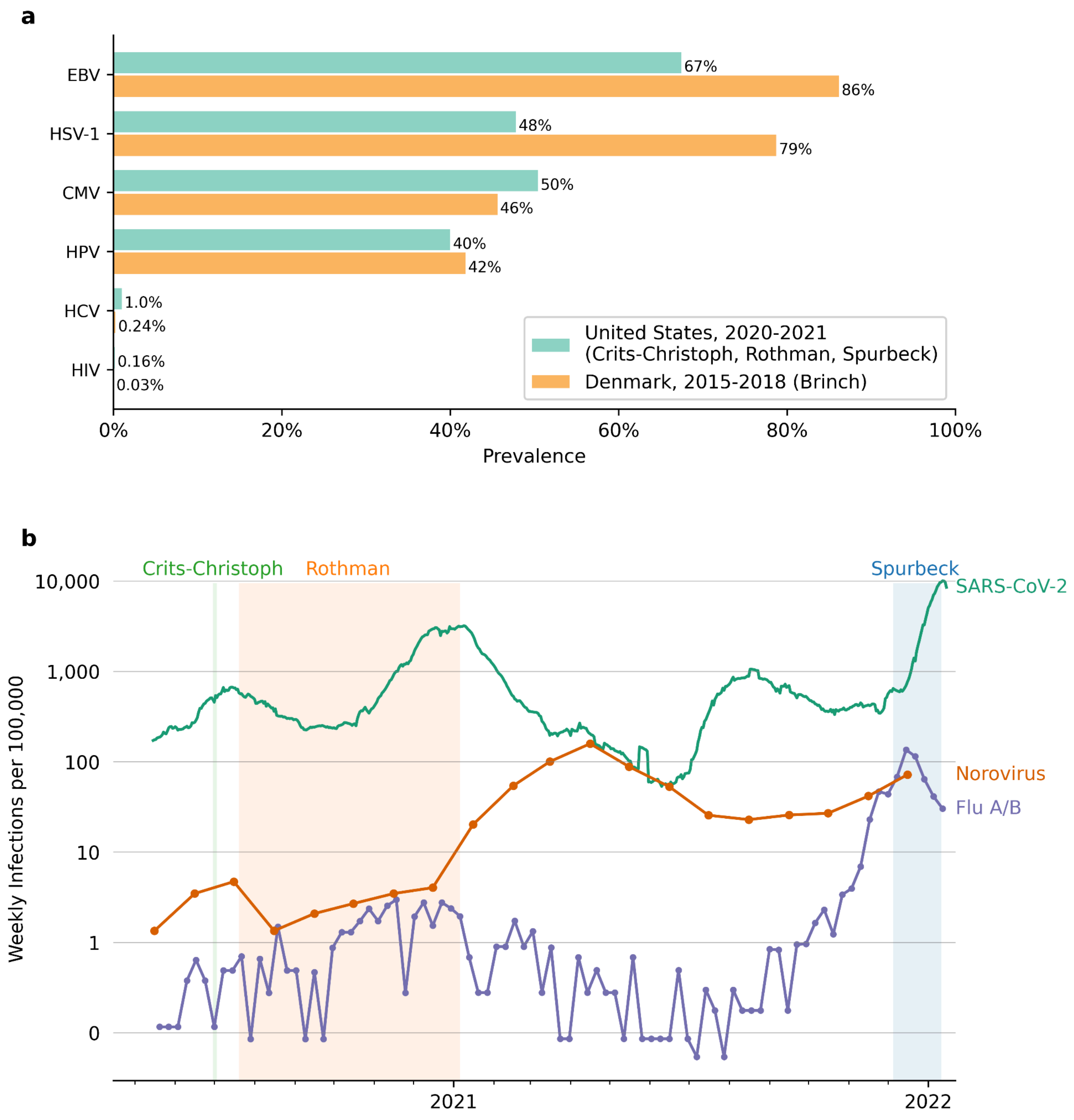


**Figure S1: Prevalence and incidence of human-infecting viruses. (a)** Prevalence of chronic-infecting viruses in the United States 2020-2021 (green), and Denmark 2015-2018 (orange) across samples for each included study. **(b)** Incidence of acute-infecting viruses in the United States, May 2020 to January 2022. SARS-CoV-2 incidence rates (dark red) are a population-weighted average of county-level daily incidences for all counties covered by target metagenomic studies. Influenza incidence rates are population-weighted averages of state-level weekly incidences for the states covered by target metagenomic studies (Ohio and California). Norovirus weekly incidence rates are based on nation-level, monthly data. No estimates for acute-infecting viruses were created for Denmark, as RNA viruses were expected to show zero read counts in DNA sequencing data. HIV estimate only applies to individuals not under treatment. Coverage of studies only shown for unenriched samples.

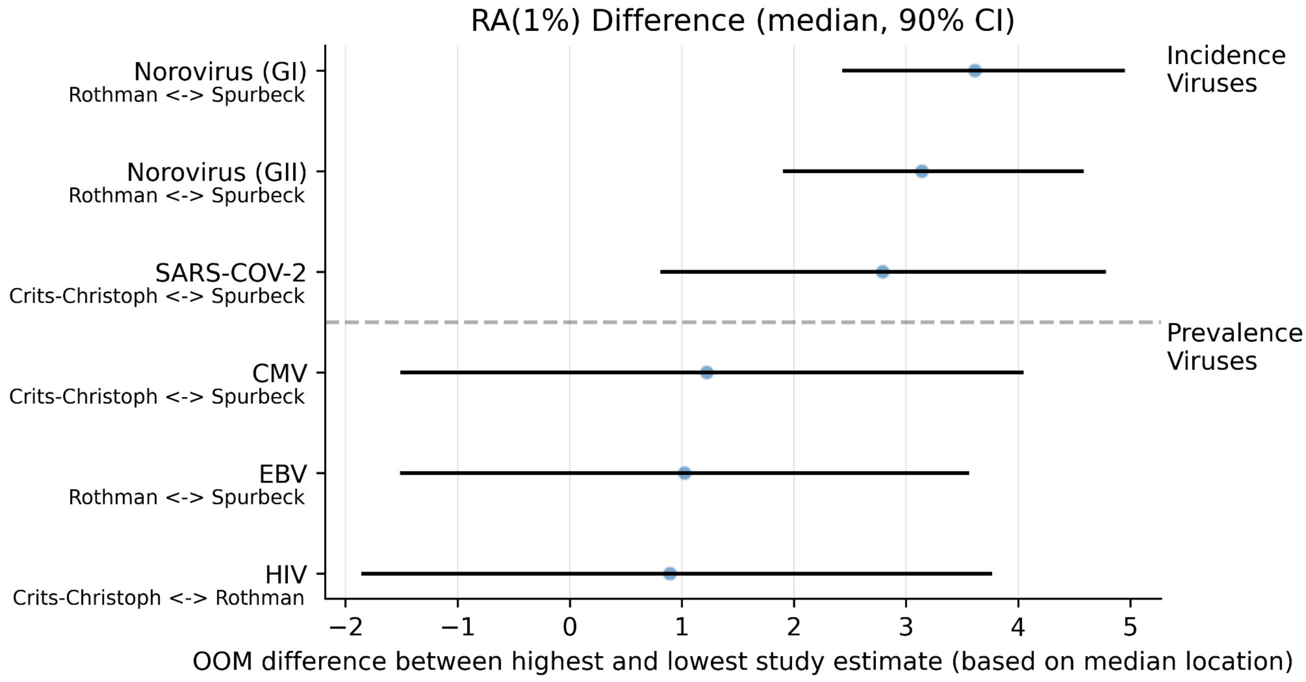


**Figure S2: Inter-study variability in RA(1%).** Displayed as the difference between the study with the highest median $RA(1\%)$ and the study with the lowest median $RA(1\%)$. The bars represent the difference between the two studies' median; the range spans i) the high-end of the lower estimate divided by the low-end of the high estimate and ii) the low-end of the lower estimate divided by the high-end of the higher estimate. Differences are displayed on a log-scale.

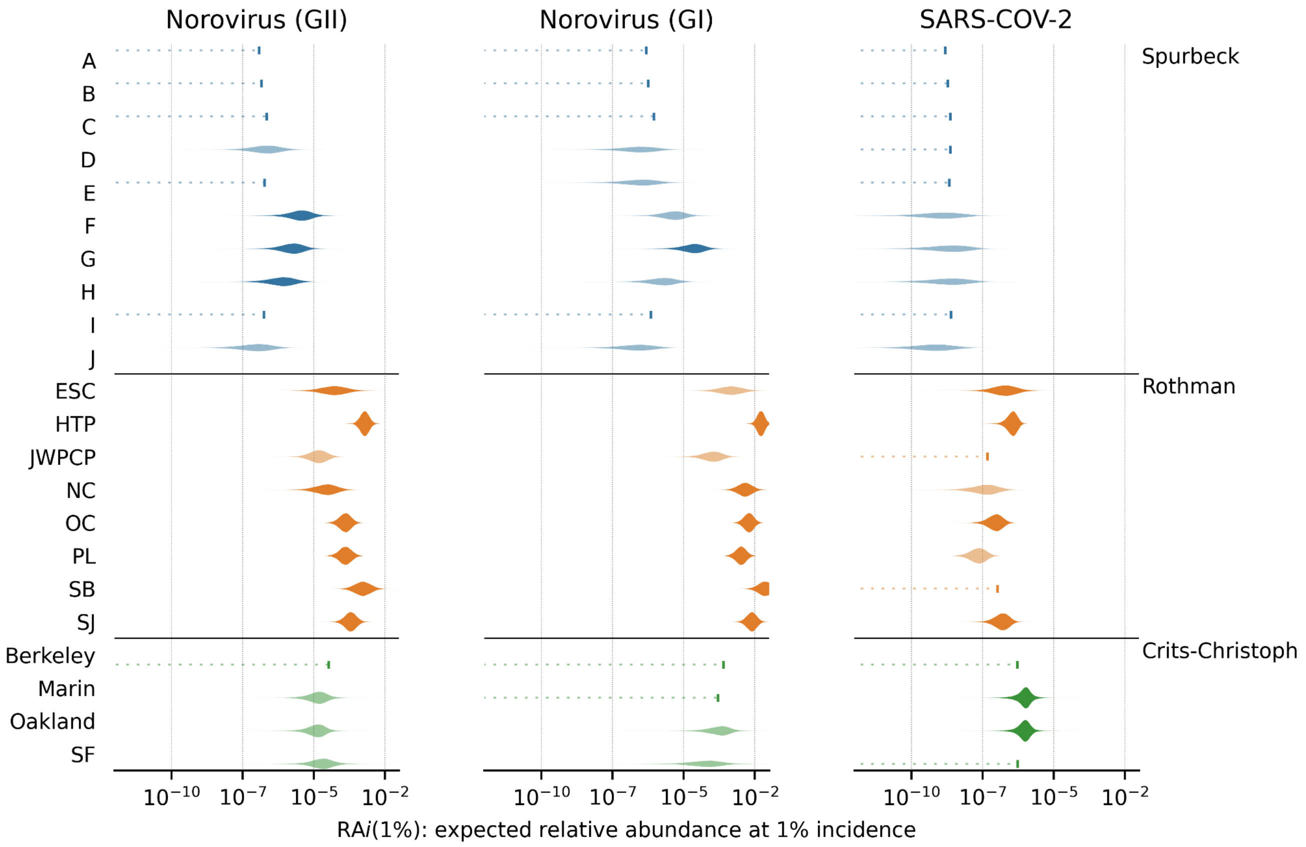


**Figure S3. Location-level relative abundance (RA) estimates for selected viruses.** Each violin represents the posterior predictive distribution of our Bayesian model for a specific study, location, and virus. Transparent violins indicate predictions made with <10 reads mapping to the corresponding virus; dashed lines indicate predictions made with zero mapped reads, with the rightmost bar indicating the upper 95th % bound.

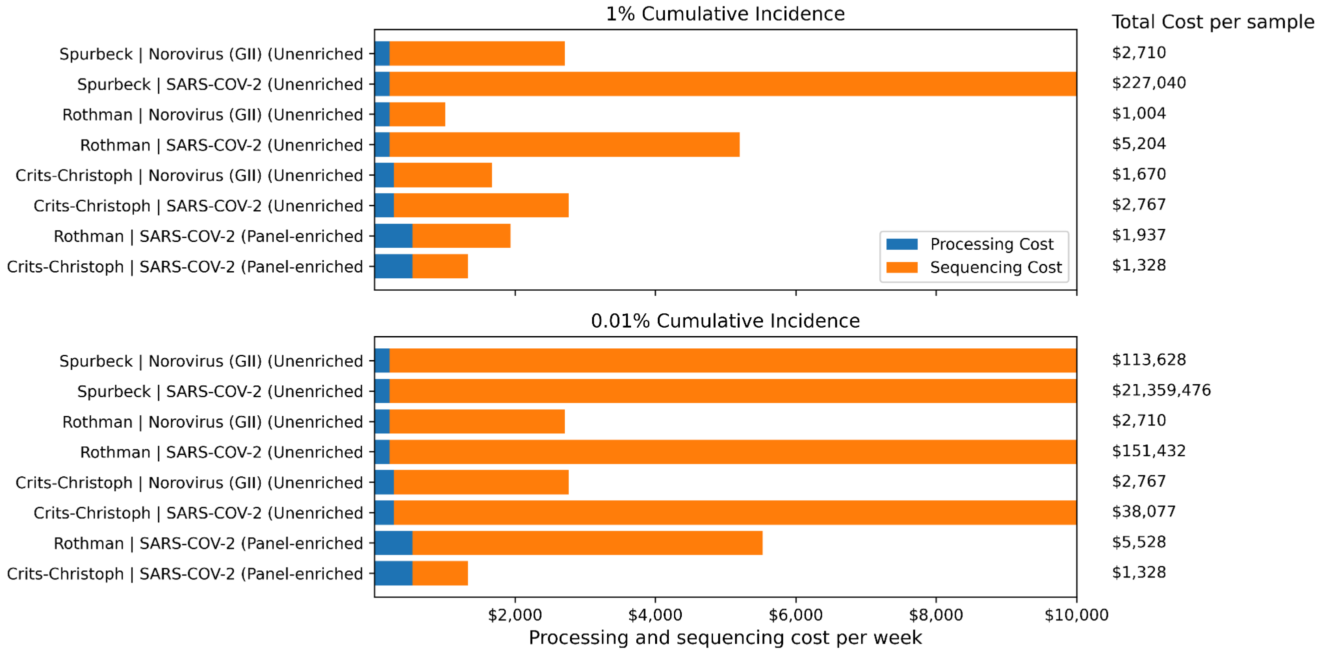


**Figure S4: Per-sample processing and sequencing cost.** Sources for processing and sequencing costs are given in appendix 6, p 25.

#### Appendix 2: Supplementary tables

| Study | Relative abundance (fraction of all reads) | |
|---|---|---|
| | All viruses | Human-infecting viruses |
| Brinch 2020 | $3{\cdot}06 \times 10^{-3}$ (1 in 327) | $3{\cdot}45 \times 10^{-6}$ (1 in 290185) |
| Spurbeck 2023 | $6{\cdot}3 \times 10^{-5}$ (1 in 15871) | $2{\cdot}52 \times 10^{-6}$ (1 in 397121) |
| Crits-Christoph 2023 Panel-enriched | $3{\cdot}58 \times 10^{-2}$ (1 in 27) | $1{\cdot}31 \times 10^{-2}$ (1 in 76) |
| Crits-Christoph 2023 Unenriched | $1{\cdot}75 \times 10^{-3}$ (1 in 571) | $1{\cdot}12 \times 10^{-5}$ (1 in 89581) |
| Rothman 2021 Unenriched | $1{\cdot}97 \times 10^{-2}$ (1 in 50) | $3{\cdot}45 \times 10^{-6}$ (1 in 290049) |
| Rothman 2021 Panel-enriched | $1{\cdot}04 \times 10^{-2}$ (1 in 95) | $9{\cdot}61 \times 10^{-6}$ (1 in 104023) |
| All studies | $6{\cdot}75 \times 10^{-3}$ (1 in 148) | $6{\cdot}53 \times 10^{-6}$ (1 in 153128) |

**Table S1: Median relative abundance of all viruses (left) and human-infecting viruses (right) in all studies included in Figure 1.** The "all studies" row gives the geometric mean across the median values for each study.

| Study | Bioprojects |
|---|---|
| Brinch 2020 | PRJEB13832, PRJEB34633 |
| Crits-Christoph 2021 | PRJNA661613 |
| Rothman 2021 | PRJNA729801 |
| Spurbeck 2023 | PRJNA924011 |

**Table S2: Bioproject IDs for all included studies.**

| Study | Sequencing Type | Country | Non-enriched Read Pairs | Enriched Read Pairs | Reference |
|---|---|---|---|---|---|
| Brinch et al. (2020) | DNA | Denmark | 4B | NA | (1) |
| Crits-Christoph et al. (2021) | RNA | United States | 300M | 8M | (2) |
| Rothman et al. (2021) | RNA | United States | 700M | 5B | (3) |
| Spurbeck et al. (2023) | RNA | United States | 2B | NA | (4) |

**Table S3: Metadata for all included studies.**

| Virus | Incidence/Prevalence | Exclusion Criterion |
|---|---|---|
| SARS-CoV-2 | Incidence | Not excluded |
| Influenza A&B Virus | Incidence | Not excluded |
| Norovirus | Incidence | Not excluded |
| Respiratory Syncytial Virus | Incidence | Heavily suppressed during the coverage period of RNA studies |
| Rhinovirus | Incidence | No precise public health data available |
| Enteric Adenovirus | Incidence | No precise public health data available |
| Metapneumovirus | Incidence | No precise public health data available |
| Herpes Simplex Virus 1 (HSV-1) | Prevalence | Not excluded |
| Herpes Simplex Virus 2 (HSV-2) | Prevalence | Not excluded |
| Cytomegalovirus (CMV) | Prevalence | Not excluded |
| Epstein-Barr-Virus (EBV) | Prevalence | Not excluded |
| Human Immunodeficiency Virus (HIV) | Prevalence | Not excluded |
| Hepatitis B Virus (HBV) | Prevalence | Not excluded |
| Hepatitis C Virus (HCV) | Prevalence | Not excluded |
| Human Papilloma Virus (HPV) | Prevalence | Not excluded |
| Hepatitis A Virus (HAV) | Prevalence | No precise public health data available |

**Table S4:** Reasons for why no epidemiological estimates were created for Respiratory Syncytial Virus, Rhinovirus, Enteric Adenovirus, Metapneumovirus, and Hepatitis A Virus.

| Virus | Abbreviation | Incidence/ Prevalence | NCBI taxid | Underlying Data for US | Underlying Data for Denmark |
|---|---|---|---|---|---|
| Norovirus | Norovirus | Incidence | 122928 (Norovirus GI) 122929 (Norovirus GII) | Yearly incidence estimate from 2006 (5), adjusted by number of outbreaks per year (6). | No estimate |
| Severe Acute Respiratory Syndrome Coronavirus 2 | SARS-CoV-2 | Incidence | 2697049 | Confirmed and probable COVID-19 cases (7), adjusted by a CDC-provided underreporting factor (8) | No estimate |
| Influenza A&B Virus | Flu A&B | Incidence | 11320 (Influenza A) 11520 (Influenza B) | Number of positive tests, adjusted by a yearly underreporting factor (9,10) | No estimate |
| Human Immunodeficiency Virus (untreated cases) | HIV | Prevalence | 11676 | 2019 CDC estimate of HIV prevalence (11), scaled by share of patients who are not immune-suppressed (11). | 2022 Undiagnosed HIV-positive individuals in Denmark (12), scaled by share of diagnoses made in Copenhagen (12). |
| Hepatitis C Virus | HCV | Prevalence | 11103 | 2013-2016 Estimate of chronic Hep C prevalence by the CDC (13) | 2016 Estimate of total HCV cases in Copenhagen, DK (14) |
| Epstein-Barr Virus | EBV | Prevalence | 10376 | CDC EBV seroprevalence measurements from NHANES cycles 2003-2010(15), adjusted for current demographics. | 1983 EBV seroprevalence measurements, adjusted for current demographics (16). |
| Herpes Simplex Virus 1 | HSV-1 | Prevalence | 10298 | 2015 CDC estimates of HSV-1 seroprevalence among 14-49 yos. Olds (17). | Extrapolated seroprevalence data based on a 2008-2011 survey of 6627 Germans (18). |
| Cytomegalo virus | CMV | Prevalence | 10359 | US-American CMV seroprevalence from NHANES cycle 1999-2004 (19). | Estimate of Dutch seroprevalence, based on seroprevalence measurement of 6382 Dutch sera in 2006 (20). |
| Human Papillomavirus | HPV | Prevalence | 10566 | 2018 CDC HPV prevalence estimates among 15-59-year-olds (21). | HPV PCR measurements in a 2016 male Danish cohort of 2436 men (22) |

**Table S5: Human-infecting viruses included in study.**

| Virus | Study | Median | 5th Percentile | 95th Percentile |
|---|---|---|---|---|
| Influenza A | Rothman | $1{\cdot}86 \times 10^{-7}$ | $4{\cdot}41 \times 10^{-9}$ | $2{\cdot}37 \times 10^{-6}$ |
| Influenza A | Crits-Christoph | $6{\cdot}23 \times 10^{-6}$ | $8{\cdot}08 \times 10^{-8}$ | $1{\cdot}73 \times 10^{-4}$ |
| Influenza A | Spurbeck | $2{\cdot}31 \times 10^{-10}$ | $4{\cdot}02 \times 10^{-12}$ | $3{\cdot}60 \times 10^{-9}$ |
| Influenza A | Mean (geometric) | $6{\cdot}45 \times 10^{-8}$ | $1{\cdot}13 \times 10^{-9}$ | $1{\cdot}14 \times 10^{-6}$ |
| Influenza B | Rothman | $2{\cdot}76 \times 10^{-7}$ | $6{\cdot}25 \times 10^{-9}$ | $3{\cdot}44 \times 10^{-6}$ |
| Influenza B | Crits-Christoph | $7{\cdot}64 \times 10^{-6}$ | $8{\cdot}58 \times 10^{-8}$ | $1{\cdot}85 \times 10^{-4}$ |
| Influenza B | Spurbeck | $1{\cdot}15 \times 10^{-8}$ | $2{\cdot}34 \times 10^{-10}$ | $1{\cdot}80 \times 10^{-7}$ |
| Influenza B | Mean (geometric) | $2{\cdot}89 \times 10^{-7}$ | $5{\cdot}00 \times 10^{-9}$ | $4{\cdot}85 \times 10^{-6}$ |
| Norovirus (GI) | Rothman | $1{\cdot}41 \times 10^{-3}$ | $4{\cdot}92 \times 10^{-4}$ | $3{\cdot}93 \times 10^{-3}$ |
| Norovirus (GI) | Crits-Christoph | $4{\cdot}73 \times 10^{-5}$ | $3{\cdot}12 \times 10^{-6}$ | $3{\cdot}13 \times 10^{-4}$ |
| Norovirus (GI) | Spurbeck | $3{\cdot}43 \times 10^{-7}$ | $4{\cdot}56 \times 10^{-8}$ | $1{\cdot}76 \times 10^{-6}$ |
| Norovirus (GI) | Mean (geometric) | $2{\cdot}84 \times 10^{-5}$ | $4{\cdot}12 \times 10^{-6}$ | $1{\cdot}29 \times 10^{-4}$ |
| Norovirus (GII) | Rothman | $1{\cdot}15 \times 10^{-4}$ | $3{\cdot}77 \times 10^{-5}$ | $3{\cdot}28 \times 10^{-4}$ |
| Norovirus (GII) | Crits-Christoph | $1{\cdot}25 \times 10^{-5}$ | $2{\cdot}13 \times 10^{-6}$ | $4{\cdot}78 \times 10^{-5}$ |
| Norovirus (GII) | Spurbeck | $8{\cdot}27 \times 10^{-8}$ | $8{\cdot}87 \times 10^{-9}$ | $4{\cdot}57 \times 10^{-7}$ |
| Norovirus (GII) | Mean (geometric) | $4{\cdot}91 \times 10^{-6}$ | $8{\cdot}93 \times 10^{-7}$ | $1{\cdot}93 \times 10^{-5}$ |
| SARS-COV-2 | Rothman | $6{\cdot}05 \times 10^{-8}$ | $1{\cdot}53 \times 10^{-8}$ | $1{\cdot}92 \times 10^{-7}$ |
| SARS-COV-2 | Crits-Christoph | $2{\cdot}33 \times 10^{-7}$ | $1{\cdot}47 \times 10^{-8}$ | $1{\cdot}60 \times 10^{-6}$ |
| SARS-COV-2 | Spurbeck | $3{\cdot}78 \times 10^{-10}$ | $2{\cdot}75 \times 10^{-11}$ | $2{\cdot}20 \times 10^{-9}$ |
| SARS-COV-2 | Mean (geometric) | $1{\cdot}75 \times 10^{-8}$ | $1{\cdot}83 \times 10^{-9}$ | $8{\cdot}78 \times 10^{-8}$ |
| CMV | Rothman | $3{\cdot}46 \times 10^{-12}$ | $8{\cdot}79 \times 10^{-14}$ | $3{\cdot}66 \times 10^{-11}$ |
| CMV | Crits-Christoph | $3{\cdot}84 \times 10^{-11}$ | $8{\cdot}98 \times 10^{-13}$ | $5{\cdot}07 \times 10^{-10}$ |
| CMV | Spurbeck | $2{\cdot}30 \times 10^{-12}$ | $4{\cdot}74 \times 10^{-14}$ | $2{\cdot}82 \times 10^{-11}$ |
| CMV | Brinch | $8{\cdot}59 \times 10^{-13}$ | $1{\cdot}61 \times 10^{-14}$ | $1{\cdot}26 \times 10^{-11}$ |
| CMV | Mean (geometric) | $4{\cdot}03 \times 10^{-12}$ | $8{\cdot}81 \times 10^{-14}$ | $5{\cdot}06 \times 10^{-11}$ |
| EBV | Rothman | $1{\cdot}38 \times 10^{-11}$ | $5{\cdot}92 \times 10^{-13}$ | $9{\cdot}59 \times 10^{-11}$ |
| EBV | Crits-Christoph | $2{\cdot}63 \times 10^{-12}$ | $4{\cdot}12 \times 10^{-14}$ | $8{\cdot}45 \times 10^{-11}$ |
| EBV | Spurbeck | $1{\cdot}30 \times 10^{-12}$ | $2{\cdot}73 \times 10^{-14}$ | $1{\cdot}86 \times 10^{-11}$ |
| EBV | Brinch | $2{\cdot}99 \times 10^{-11}$ | $7{\cdot}46 \times 10^{-12}$ | $9{\cdot}88 \times 10^{-11}$ |
| EBV | Mean (geometric) | $6{\cdot}13 \times 10^{-12}$ | $2{\cdot}66 \times 10^{-13}$ | $6{\cdot}22 \times 10^{-11}$ |
| HCV | Rothman | $8{\cdot}30 \times 10^{-11}$ | $1{\cdot}64 \times 10^{-12}$ | $1{\cdot}19 \times 10^{-9}$ |
| HCV | Crits-Christoph | $2{\cdot}77 \times 10^{-10}$ | $2{\cdot}86 \times 10^{-12}$ | $6{\cdot}74 \times 10^{-9}$ |
| HCV | Spurbeck | $5{\cdot}62 \times 10^{-11}$ | $1{\cdot}21 \times 10^{-12}$ | $7{\cdot}48 \times 10^{-10}$ |
| HCV | Brinch | $7{\cdot}81 \times 10^{-11}$ | $1{\cdot}22 \times 10^{-12}$ | $1{\cdot}74 \times 10^{-9}$ |
| HCV | Mean (geometric) | $1{\cdot}00 \times 10^{-10}$ | $1{\cdot}62 \times 10^{-12}$ | $1{\cdot}80 \times 10^{-9}$ |
| HIV | Rothman | $9{\cdot}94 \times 10^{-10}$ | $1{\cdot}74 \times 10^{-11}$ | $1{\cdot}34 \times 10^{-8}$ |
| HIV | Crits-Christoph | $7{\cdot}79 \times 10^{-9}$ | $1{\cdot}93 \times 10^{-10}$ | $9{\cdot}79 \times 10^{-8}$ |
| HIV | Spurbeck | $3{\cdot}72 \times 10^{-10}$ | $6{\cdot}97 \times 10^{-12}$ | $5{\cdot}31 \times 10^{-9}$ |

| | | | | |
|---|---|---|---|---|
| HIV | Brinch | $1{\cdot}70 \times 10^{-9}$ | $3{\cdot}23 \times 10^{-11}$ | $2{\cdot}31 \times 10^{-8}$ |
| HIV | Mean (geometric) | $1{\cdot}49 \times 10^{-9}$ | $2{\cdot}95 \times 10^{-11}$ | $2{\cdot}00 \times 10^{-8}$ |
| HPV | Rothman | $2{\cdot}16 \times 10^{-12}$ | $3{\cdot}80 \times 10^{-14}$ | $3{\cdot}21 \times 10^{-11}$ |
| HPV | Crits-Christoph | $7{\cdot}82 \times 10^{-12}$ | $7{\cdot}63 \times 10^{-14}$ | $1{\cdot}72 \times 10^{-10}$ |
| HPV | Spurbeck | $1{\cdot}44 \times 10^{-12}$ | $2{\cdot}59 \times 10^{-14}$ | $2{\cdot}03 \times 10^{-11}$ |
| HPV | Brinch | $8{\cdot}78 \times 10^{-13}$ | $1{\cdot}71 \times 10^{-14}$ | $2{\cdot}12 \times 10^{-11}$ |
| HPV | Mean (geometric) | $2{\cdot}15 \times 10^{-12}$ | $3{\cdot}36 \times 10^{-14}$ | $3{\cdot}93 \times 10^{-11}$ |
| HSV-1 | Rothman | $5{\cdot}11 \times 10^{-10}$ | $2{\cdot}16 \times 10^{-10}$ | $9{\cdot}89 \times 10^{-10}$ |
| HSV-1 | Crits-Christoph | $5{\cdot}88 \times 10^{-12}$ | $6{\cdot}56 \times 10^{-14}$ | $1{\cdot}48 \times 10^{-10}$ |
| HSV-1 | Spurbeck | $1{\cdot}11 \times 10^{-12}$ | $1{\cdot}79 \times 10^{-14}$ | $1{\cdot}62 \times 10^{-11}$ |
| HSV-1 | Brinch | $5{\cdot}87 \times 10^{-11}$ | $1{\cdot}87 \times 10^{-11}$ | $1{\cdot}79 \times 10^{-10}$ |
| HSV-1 | Mean (geometric) | $2{\cdot}10 \times 10^{-11}$ | $1{\cdot}47 \times 10^{-12}$ | $1{\cdot}44 \times 10^{-10}$ |

**Table S6: RA(1%) estimates for human-infecting viruses in unenriched sequencing data**. “Mean (geometric)” entries show the simple geometric mean across the three point estimates for individual studies.

| Virus | Study | Median | 5th Percentile | 95th Percentile | Ratio (enriched:unenriched, median) |
|---|---|---|---|---|---|
| Influenza A | Rothman Unenriched | $1·86 \times 10^{-7}$ | $4·41 \times 10^{-9}$ | $2·37 \times 10^{-6}$ | |
| Influenza A | Rothman Panel-enriched | $1·63 \times 10^{-7}$ | $7·86 \times 10^{-9}$ | $2·35 \times 10^{-6}$ | $8.73 \times 10^{-1}$ |
| Influenza A | Crits-Christoph Unenriched | $6·23 \times 10^{-6}$ | $8·08 \times 10^{-8}$ | $1·73 \times 10^{-4}$ | |
| Influenza A | Crits-Christoph Panel-enriched | $6·71 \times 10^{-4}$ | $7·58 \times 10^{-6}$ | $1·88 \times 10^{-2}$ | 107·83 |
| Influenza B | Rothman Unenriched | $2·76 \times 10^{-7}$ | $6·25 \times 10^{-9}$ | $3·44 \times 10^{-6}$ | |
| Influenza B | Rothman Panel-enriched | $7·89 \times 10^{-8}$ | $1·59 \times 10^{-9}$ | $1·23 \times 10^{-6}$ | $2·86 \times 10^{-1}$ |
| Influenza B | Crits-Christoph Unenriched | $7·64 \times 10^{-6}$ | $8·58 \times 10^{-8}$ | $1·85 \times 10^{-4}$ | |
| Influenza B | Crits-Christoph Panel-enriched | $7·84 \times 10^{-4}$ | $9·31 \times 10^{-6}$ | $2·26 \times 10^{-2}$ | 102·63 |
| Norovirus (GI) | Rothman Unenriched | $1·41 \times 10^{-3}$ | $4·92 \times 10^{-4}$ | $3·93 \times 10^{-3}$ | |
| Norovirus (GI) | Rothman Panel-enriched | $2·22 \times 10^{-4}$ | $4·55 \times 10^{-5}$ | $1·22 \times 10^{-3}$ | $1·58 \times 10^{-1}$ |
| Norovirus (GI) | Crits-Christoph Unenriched | $4·73 \times 10^{-5}$ | $3·12 \times 10^{-6}$ | $3·13 \times 10^{-4}$ | |
| Norovirus (GI) | Crits-Christoph Panel-enriched | $5·76 \times 10^{-4}$ | $1·25 \times 10^{-5}$ | $8·85 \times 10^{-3}$ | 12·17 |
| Norovirus (GII) | Rothman Unenriched | $1·15 \times 10^{-4}$ | $3·77 \times 10^{-5}$ | $3·28 \times 10^{-4}$ | |
| Norovirus (GII) | Rothman Panel-enriched | $2·40 \times 10^{-5}$ | $6·11 \times 10^{-6}$ | $1·01 \times 10^{-4}$ | $2·09 \times 10^{-1}$ |
| Norovirus (GII) | Crits-Christoph Unenriched | $1·25 \times 10^{-5}$ | $2·13 \times 10^{-6}$ | $4·78 \times 10^{-5}$ | |
| Norovirus (GII) | Crits-Christoph Panel-enriched | $3·23 \times 10^{-4}$ | $1·74 \times 10^{-5}$ | $2·83 \times 10^{-3}$ | 25·86 |
| SARS-COV-2 | Rothman Unenriched | $6·05 \times 10^{-8}$ | $1·53 \times 10^{-8}$ | $1·92 \times 10^{-7}$ | |
| SARS-COV-2 | Rothman Panel-enriched | $4·11 \times 10^{-6}$ | $1·11 \times 10^{-6}$ | $1·66 \times 10^{-5}$ | 67·94 |
| SARS-COV-2 | Crits-Christoph Unenriched | $2·33 \times 10^{-7}$ | $1·47 \times 10^{-8}$ | $1·60 \times 10^{-6}$ | |
| SARS-COV-2 | Crits-Christoph Panel-enriched | $1·66 \times 10^{-1}$ | $2·10 \times 10^{-2}$ | $5·84 \times 10^{-1}$ | 714067·43 |
| CMV | Rothman Unenriched | $3·46 \times 10^{-12}$ | $8·79 \times 10^{-14}$ | $3·66 \times 10^{-11}$ | |
| CMV | Rothman Panel-enriched | $1·59 \times 10^{-12}$ | $1·05 \times 10^{-13}$ | $1·04 \times 10^{-11}$ | $4·59 \times 10^{-1}$ |
| CMV | Crits-Christoph Unenriched | $3·84 \times 10^{-11}$ | $8·98 \times 10^{-13}$ | $5·07 \times 10^{-10}$ | |
| CMV | Crits-Christoph Panel-enriched | $6·02 \times 10^{-10}$ | $6·76 \times 10^{-12}$ | $1·73 \times 10^{-8}$ | 15·66 |
| EBV | Rothman Unenriched | $1·38 \times 10^{-11}$ | $5·92 \times 10^{-13}$ | $9·59 \times 10^{-11}$ | |
| EBV | Rothman Panel-enriched | $5·99 \times 10^{-12}$ | $2·77 \times 10^{-13}$ | $3·95 \times 10^{-11}$ | $4·35 \times 10^{-1}$ |
| EBV | Crits-Christoph Unenriched | $2·63 \times 10^{-12}$ | $4·12 \times 10^{-14}$ | $8·45 \times 10^{-11}$ | |
| EBV | Crits-Christoph Panel-enriched | $4·18 \times 10^{-10}$ | $4·49 \times 10^{-12}$ | $1·26 \times 10^{-8}$ | 159·14 |
| HCV | Rothman Unenriched | $8·30 \times 10^{-11}$ | $1·64 \times 10^{-12}$ | $1·19 \times 10^{-9}$ | |
| HCV | Rothman Panel-enriched | $1·57 \times 10^{-11}$ | $4·25 \times 10^{-13}$ | $2·46 \times 10^{-10}$ | $1·89 \times 10^{-1}$ |
| HCV | Crits-Christoph Unenriched | $2·77 \times 10^{-10}$ | $2·86 \times 10^{-12}$ | $6·74 \times 10^{-9}$ | |
| HCV | Crits-Christoph Panel-enriched | $2·77 \times 10^{-8}$ | $3·50 \times 10^{-10}$ | $7·99 \times 10^{-7}$ | 100·04 |
| HIV | Rothman Unenriched | $9·94 \times 10^{-10}$ | $1·74 \times 10^{-11}$ | $1·34 \times 10^{-8}$ | |
| HIV | Rothman Panel-enriched | $2·54 \times 10^{-9}$ | $3·54 \times 10^{-10}$ | $9·47 \times 10^{-9}$ | 2·55 |
| HIV | Crits-Christoph Unenriched | $7·79 \times 10^{-9}$ | $1·93 \times 10^{-10}$ | $9·79 \times 10^{-8}$ | |
| HIV | Crits-Christoph Panel-enriched | $1·76 \times 10^{-7}$ | $2·15 \times 10^{-9}$ | $5·29 \times 10^{-6}$ | 22·64 |
| HPV | Rothman Unenriched | $2·16 \times 10^{-12}$ | $3·80 \times 10^{-14}$ | $3·21 \times 10^{-11}$ | |
| HPV | Rothman Panel-enriched | $4·78 \times 10^{-13}$ | $1·00 \times 10^{-14}$ | $6·10 \times 10^{-12}$ | $2·21 \times 10^{-1}$ |

| | | | | | |
|---|---|---|---|---|---|
| HPV | Crits-Christoph Unenriched | $7{\cdot}82 \times 10^{-12}$ | $7{\cdot}63 \times 10^{-14}$ | $1{\cdot}72 \times 10^{-10}$ | |
| HPV | Crits-Christoph Panel-enriched | $6{\cdot}36 \times 10^{-10}$ | $1{\cdot}00 \times 10^{-11}$ | $1{\cdot}99 \times 10^{-8}$ | 81·26 |
| HSV-1 | Rothman Unenriched | $5{\cdot}11 \times 10^{-10}$ | $2{\cdot}16 \times 10^{-10}$ | $9{\cdot}89 \times 10^{-10}$ | |
| HSV-1 | Rothman Panel-enriched | $7{\cdot}90 \times 10^{-11}$ | $2{\cdot}95 \times 10^{-11}$ | $1{\cdot}87 \times 10^{-10}$ | $1{\cdot}55 \times 10^{-1}$ |
| HSV-1 | Crits-Christoph Unenriched | $5{\cdot}88 \times 10^{-12}$ | $6{\cdot}56 \times 10^{-14}$ | $1{\cdot}48 \times 10^{-10}$ | |
| HSV-1 | Crits-Christoph Panel-enriched | $6{\cdot}05 \times 10^{-10}$ | $6{\cdot}13 \times 10^{-12}$ | $1{\cdot}73 \times 10^{-8}$ | 102·81 |

**Table S7: Comparison of RA(1%) between hybridization-capture and unenriched samples.** The “Ratio” column gives the simple ratio between the median RA(1%) estimates for enriched and unenriched samples.

| Virus | Study | Median | 5th Percentile | 95th Percentile |
|---|---|---|---|---|
| Norovirus (GII) | Crits-Christoph | $8{\cdot}01 \times 10^{6}$ | $2{\cdot}09 \times 10^{6}$ | $4{\cdot}69 \times 10^{7}$ |
| Norovirus (GII) | Rothman | $8{\cdot}71 \times 10^{5}$ | $3{\cdot}05 \times 10^{5}$ | $2{\cdot}65 \times 10^{6}$ |
| Norovirus (GII) | Spurbeck | $1{\cdot}21 \times 10^{9}$ | $2{\cdot}19 \times 10^{8}$ | $1{\cdot}13 \times 10^{10}$ |
| Norovirus (GII) | Mean (geometric) | $2{\cdot}04 \times 10^{7}$ | $5{\cdot}19 \times 10^{6}$ | $1{\cdot}12 \times 10^{8}$ |
| SARS-COV-2 | Crits-Christoph | $4{\cdot}29 \times 10^{8}$ | $6{\cdot}24 \times 10^{7}$ | $6{\cdot}82 \times 10^{9}$ |
| SARS-COV-2 | Rothman | $1{\cdot}65 \times 10^{9}$ | $5{\cdot}21 \times 10^{8}$ | $6{\cdot}54 \times 10^{9}$ |
| SARS-COV-2 | Spurbeck | $2{\cdot}64 \times 10^{11}$ | $4{\cdot}54 \times 10^{10}$ | $3{\cdot}64 \times 10^{12}$ |
| SARS-COV-2 | Mean (geometric) | $5{\cdot}72 \times 10^{9}$ | $1{\cdot}14 \times 10^{9}$ | $5{\cdot}45 \times 10^{10}$ |

**Table S8: Weekly sequencing required for detection at 1% cumulative incidence and a detection threshold of 100 reads.** Changing the detection threshold alters the required read depth proportionally (Equation A3.6).

| Virus | Study | Median | 5th Percentile | 95th Percentile | Detection Threshold |
|---|---|---|---|---|---|
| Norovirus (GII) | Rothman Panel-enriched | $4{\cdot}17 \times 10^{6}$ | $9{\cdot}91 \times 10^{5}$ | $1{\cdot}64 \times 10^{7}$ | 100 |
| Norovirus (GII) | Crits-Christoph Panel-enriched | $3{\cdot}10 \times 10^{5}$ | $3{\cdot}53 \times 10^{4}$ | $5{\cdot}76 \times 10^{6}$ | 100 |
| Norovirus (GII) | Mean (geometric) | $1{\cdot}14 \times 10^{6}$ | $1{\cdot}87 \times 10^{5}$ | $9{\cdot}71 \times 10^{6}$ | 100 |
| SARS-COV-2 | Rothman Panel-enriched | $2{\cdot}43 \times 10^{7}$ | $6{\cdot}01 \times 10^{6}$ | $9{\cdot}03 \times 10^{7}$ | 100 |
| SARS-COV-2 | Crits-Christoph Panel-enriched | $6{\cdot}01 \times 10^{2}$ | $1{\cdot}71 \times 10^{2}$ | $4{\cdot}77 \times 10^{3}$ | 100 |
| SARS-COV-2 | Mean (geometric) | $1{\cdot}21 \times 10^{5}$ | $3{\cdot}21 \times 10^{4}$ | $6{\cdot}57 \times 10^{5}$ | 100 |
| Influenza A | Rothman Panel-enriched | $6{\cdot}15 \times 10^{8}$ | $4{\cdot}26 \times 10^{7}$ | $1{\cdot}27 \times 10^{10}$ | 100 |
| Influenza A | Crits-Christoph Panel-enriched | $1{\cdot}49 \times 10^{5}$ | $5{\cdot}33 \times 10^{3}$ | $1{\cdot}32 \times 10^{7}$ | 100 |
| Influenza A | Mean (geometric) | $9{\cdot}57 \times 10^{6}$ | $4{\cdot}76 \times 10^{5}$ | $4{\cdot}10 \times 10^{8}$ | 100 |

**Table S9: Weekly sequencing required for detection at 1% cumulative incidence for hybridization capture samples.** Changing the detection threshold alters the required read depth proportionally (Equation A3.6).

| Virus | Enrichment | Cumulative incidence | Study | Processing cost (per sample) $c_j^{proc}$ | Sequencing cost (per sample) $c_j^{seq}$ | Total cost (per sample) $c_j$ | Total cost (per year) $C_j$ | Sequencing depth $n_j$ | Sequencing platform $k$ |
|---|---|---|---|---|---|---|---|---|---|
| SARS-COV-2 | Panel-enriched | 1.00% | Crits-Christoph | $540 | $788 | $1,328 | $69,056 | $6.01 \times 10^2$ | MiSeq |
| SARS-COV-2 | Panel-enriched | 1.00% | Rothman | $540 | $1,397 | $1,937 | $100,724 | $2.43 \times 10^7$ | NextSeq |
| SARS-COV-2 | Panel-enriched | 1.00% | Overall (geomean) | $540 | $1,049 | $1,603 | $83,400 | $1.21 \times 10^5$ | N/A |
| SARS-COV-2 | Panel-enriched | 0.01% | Crits-Christoph | $540 | $788 | $1,328 | $69,056 | $6.01 \times 10^4$ | MiSeq |
| SARS-COV-2 | Panel-enriched | 0.01% | Rothman | $540 | $4,988 | $5,528 | $287,456 | $2.43 \times 10^9$ | NovaSeq (lane) |
| SARS-COV-2 | Panel-enriched | 0.01% | Overall (geomean) | $540 | $1,982 | $2,709 | $140,892 | $1.21 \times 10^7$ | N/A |
| SARS-COV-2 | Unenriched | 1.00% | Crits-Christoph | $273 | $2,494 | $2,767 | $143,884 | $4.29 \times 10^8$ | NovaSeq (lane) |
| SARS-COV-2 | Unenriched | 1.00% | Rothman | $216 | $4,988 | $5,204 | $270,608 | $1.65 \times 10^9$ | NovaSeq (lane) |
| SARS-COV-2 | Unenriched | 1.00% | Spurbeck | $216 | $226,824 | $227,040 | $11,806,080 | $2.64 \times 10^{11}$ | NovaSeq (cell) |
| SARS-COV-2 | Unenriched | 1.00% | Overall (geomean) | $233 | $14,130 | $14,841 | $771,767 | $5.72 \times 10^9$ | N/A |
| SARS-COV-2 | Unenriched | 0.01% | Crits-Christoph | $273 | $37,804 | $38,077 | $1,980,004 | $4.29 \times 10^{10}$ | NovaSeq (cell) |
| SARS-COV-2 | Unenriched | 0.01% | Rothman | $216 | $151,216 | $151,432 | $7,874,464 | $1.65 \times 10^{11}$ | NovaSeq (cell) |
| SARS-COV-2 | Unenriched | 0.01% | Spurbeck | $216 | $21,359,260 | $21,359,476 | $1,110,692,752 | $2.64 \times 10^{13}$ | NovaSeq (cell) |
| SARS-COV-2 | Unenriched | 0.01% | Overall (geomean) | $233 | $496,105 | $497,535 | $25,871,821 | $5.72 \times 10^{11}$ | N/A |
| Norovirus (GII) | Unenriched | 1.00% | Crits-Christoph | $273 | $1,397 | $1,670 | $86,840 | $8.01 \times 10^6$ | NextSeq |
| Norovirus (GII) | Unenriched | 1.00% | Rothman | $216 | $788 | $1,004 | $52,208 | $8.71 \times 10^5$ | MiSeq |
| Norovirus (GII) | Unenriched | 1.00% | Spurbeck | $216 | $2,494 | $2,710 | $140,920 | $1.21 \times 10^9$ | NovaSeq (lane) |
| Norovirus (GII) | Unenriched | 1.00% | Overall (geomean) | $233 | $1,400 | $1,656 | $86,127 | $2.04 \times 10^7$ | N/A |
| Norovirus (GII) | Unenriched | 0.01% | Crits-Christoph | $273 | $2,494 | $2,767 | $143,884 | $8.01 \times 10^8$ | NovaSeq (lane) |
| Norovirus (GII) | Unenriched | 0.01% | Rothman | $216 | $2,494 | $2,710 | $140,920 | $8.71 \times 10^7$ | NovaSeq (lane) |
| Norovirus (GII) | Unenriched | 0.01% | Spurbeck | $216 | $113,412 | $113,628 | $5,908,656 | $1.21 \times 10^{11}$ | NovaSeq (cell) |
| Norovirus (GII) | Unenriched | 0.01% | Overall (geomean) | $233 | $8,901 | $9,480 | $492,974 | $2.04 \times 10^9$ | N/A |

**Table S10: Estimated sequencing costs and depths for detection of SARS-CoV-2 and Norovirus (GII) at 1% and 0·01% cumulative incidence.** Costs are shown per sample and annually, assuming weekly sampling. Estimates are provided for different studies (Crits-Christoph, Rothman, Spurbeck) and sample types (panel-enriched, unenriched). Sequencing platform selection (NovaSeq (full flow cell), NovaSeq (flow cell lane), NextSeq, or MiSeq) was based on the most cost-effective option for the required sequencing depth. Costs include library preparation, and where applicable, panel enrichment or RNA depletion. A detection threshold of 100 reads was assumed for all scenarios. Equations for cost variables are given in Appendix 6.

| Pathogen | Enrichment | Study | P2RA at 0.01 | P2RA at 0.1 (% Decrease) | P2RA at 1 (% Decrease) |
|---|---|---|---|---|---|
| Influenza A | enriched | Crits-Christoph et al. 2021 | $5.55 \times 10^{-3}$ | $7.18 \times 10^{-4}$ (87.1%) | $7.39 \times 10^{-5}$ (98.7%) |
| Influenza A | enriched | Rothman et al. 2021 | $1.14 \times 10^{-7}$ | $1.53 \times 10^{-7}$ (-34.5%) | $8.21 \times 10^{-8}$ (28.1%) |
| Influenza A | unenriched | Crits-Christoph et al. 2021 | $7.19 \times 10^{-5}$ | $5.30 \times 10^{-6}$ (92.6%) | $7.19 \times 10^{-7}$ (99.0%) |
| Influenza A | unenriched | Rothman et al. 2021 | $2.43 \times 10^{-7}$ | $2.20 \times 10^{-7}$ (9.4%) | $1.21 \times 10^{-7}$ (50.1%) |
| Influenza A | unenriched | Spurbeck et al. 2023 | $3.37 \times 10^{-10}$ | $3.37 \times 10^{-10}$ (0.0%) | $3.37 \times 10^{-10}$ (0.0%) |
| Influenza B | enriched | Crits-Christoph et al. 2021 | $7.96 \times 10^{-3}$ | $7.16 \times 10^{-4}$ (91.0%) | $7.28 \times 10^{-5}$ (99.1%) |
| Influenza B | enriched | Rothman et al. 2021 | $1.02 \times 10^{-7}$ | $6.00 \times 10^{-8}$ (41.0%) | $2.94 \times 10^{-8}$ (71.1%) |
| Influenza B | unenriched | Crits-Christoph et al. 2021 | $6.42 \times 10^{-5}$ | $8.37 \times 10^{-6}$ (87.0%) | $6.42 \times 10^{-7}$ (99.0%) |
| Influenza B | unenriched | Rothman et al. 2021 | $4.34 \times 10^{-7}$ | $3.25 \times 10^{-7}$ (25.1%) | $1.53 \times 10^{-7}$ (64.8%) |
| Influenza B | unenriched | Spurbeck et al. 2023 | $1.43 \times 10^{-8}$ | $1.43 \times 10^{-8}$ (0.0%) | $1.43 \times 10^{-8}$ (0.0%) |
| Norovirus (GI) | enriched | Crits-Christoph et al. 2021 | $1.30 \times 10^{-4}$ | $1.30 \times 10^{-4}$ (0.0%) | $1.30 \times 10^{-4}$ (0.0%) |
| Norovirus (GI) | enriched | Rothman et al. 2021 | $2.61 \times 10^{-6}$ | $2.61 \times 10^{-6}$ (0.0%) | $2.61 \times 10^{-6}$ (0.0%) |
| Norovirus (GI) | unenriched | Crits-Christoph et al. 2021 | $9.21 \times 10^{-7}$ | $9.21 \times 10^{-7}$ (0.0%) | $9.21 \times 10^{-7}$ (0.0%) |
| Norovirus (GI) | unenriched | Rothman et al. 2021 | $1.88 \times 10^{-4}$ | $1.88 \times 10^{-4}$ (0.0%) | $1.88 \times 10^{-4}$ (0.0%) |
| Norovirus (GI) | unenriched | Spurbeck et al. 2023 | $1.47 \times 10^{-7}$ | $1.47 \times 10^{-7}$ (0.0%) | $1.47 \times 10^{-7}$ (0.0%) |
| Norovirus (GII) | enriched | Crits-Christoph et al. 2021 | $3.49 \times 10^{-5}$ | $3.49 \times 10^{-5}$ (0.0%) | $3.49 \times 10^{-5}$ (0.0%) |
| Norovirus (GII) | enriched | Rothman et al. 2021 | $2.84 \times 10^{-6}$ | $2.84 \times 10^{-6}$ (0.0%) | $2.84 \times 10^{-6}$ (0.0%) |
| Norovirus (GII) | unenriched | Crits-Christoph et al. 2021 | $1.21 \times 10^{-6}$ | $1.21 \times 10^{-6}$ (0.0%) | $1.21 \times 10^{-6}$ (0.0%) |
| Norovirus (GII) | unenriched | Rothman et al. 2021 | $4.17 \times 10^{-5}$ | $4.17 \times 10^{-5}$ (0.0%) | $4.17 \times 10^{-5}$ (0.0%) |
| Norovirus (GII) | unenriched | Spurbeck et al. 2023 | $4.42 \times 10^{-8}$ | $4.42 \times 10^{-8}$ (0.0%) | $4.42 \times 10^{-8}$ (0.0%) |

**Table S11: Effect on varying pseudocounts (total positive cases in area over one week) on final RA(1%) values for acute-infecting pathogens.** Columns both show the final overlap RA(1%) for each study and pathogen, and for the decrease in RA(1%) as the pseudocounts range from 0.01 to 0.1 to 1. Pseudocounts are only applied whenever a pathogen showed zero cases for a given week and region.

#### Appendix 3: Statistical inference and required sequencing depth

##### Estimating the PH-to-RA conversion factor and RA(1%)

The goal of our model was predicting the relative abundance (RA) of a given virus (henceforth the "focal virus") in W-MGS data as a function of its incidence or prevalence (henceforth "public health (PH) predictor") in the population.

We define the relative abundance of a pathogen in a W-MGS sample as the number of high-quality distinct (non-duplicate) reads divided by the total number of reads sequenced. We make the natural assumption that the expected relative abundance for a given pathogen in a given study, conditional on the PH predictor $x$, increases in proportion to $x$ at small values, saturating at $RA \approx 1$. More precisely, we define a proportionality constant $B$ such that

$$E[RA] = \frac{B \cdot x}{1 + B \cdot x} = odds^{-1}(B \cdot x) \qquad \text{(A3.1)}$$

$$\approx B \cdot x \text{ for } B \cdot x \ll 1,$$

where $odds^{-1}$ is the inverse odds transformation $odds^{-1}(x) = x/(1 + x)$, which maps an odds-transformed probability $p/(1 - p)$ back to the corresponding probability $p$. We can think of $B$ as the conversion factor between the PH predictor and the pathogen's RA (up to random measurement error in the PH predictor and the RA). It accounts for a variety of factors that influence the relative abundance of the pathogen in the W-MGS measurement, including

1. the rate of shedding of the pathogen into the wastewater system relative to the rest of the human microbiome;
2. changes in relative abundance during transport through the sewer system, for example due to decay of the pathogen and growth of the background microbiome;
3. biases during sample processing and sequencing that may increase or decrease the pathogen's measured abundance relative to the background microbiome (23).
4. sequence quality and rates of PCR duplication—lower quality and/or higher duplicate rates tend to decrease the number of high-quality, distinct pathogen reads relative to the number of total reads;
5. systematic error in the PH predictor—we use the best available public health predictor for a given sampling location, yet the population the public health predictor is derived from (e.g., the entire United States for all of 2021) may systematically differ from the population contributing to the sample (e.g., Orange County on May 21, 2021).

To go from expected to observed RA, we must also account for noise in the W-MGS measurement. Random counting error in the observed number of pathogen reads naturally arises during sequencing; this error is typically represented by the Binomial or Poisson distribution. However, additional sources of error in MGS measurements add greater variation beyond what is captured by a Binomial or Poisson model ("overdispersion"). These error sources include random deviation in the actual PH metric for the catchment at the time of sampling from the PH predictor, as well as variation in the pathogen RA due to random fluctuations in shedding, decay during transport, and bias during sample processing. We account for overdispersion by multiplying $B \cdot x$ by a lognormally-distributed noise factor $f$ with scale parameter σ. Putting these considerations together, we model the number of pathogen reads $y_s$ observed in a sample $s$ that has been sequenced to a total depth of $n_s$ reads conditional on the PH predictor

$$y_s \sim Binomial(n_s,\ odds^{-1}(B \cdot x_s \cdot f_s)) \qquad \text{(A3.2)}$$

$$f_s \sim LogNormal(0, \sigma),$$

with the observed RA equal to $y_s/n_s$.

By fitting this model to observed pathogen reads $y_s$, total reads $n_s$, and PH predictor $x_s$, we can estimate the conversion factor $B$. We can further use the fitted model to understand how the pathogen reads and/or RA in a sample scales with the PH predictor $x$ that was not observed in the data. The expected RA at a value of $x$ can be computed by substituting the estimate $\hat{B}$ in for $B$ in Equation A3.1. In particular, the expected RA when the PH predictor is 1% is given by

$$RA(1\%) = odds^{-1}(B \cdot \frac{1}{100})$$

$$\approx \frac{B}{100} \text{ for } \frac{B}{100} \ll 1.$$

This equation is applied to posterior samples of $\hat{B}$ to determine the posterior distribution of the estimated $RA(1\%)$ in the main results figures.

To fit model this model to data from the various studies, we developed a Bayesian hierarchical logistic regression model using the Stan probabilistic programming language[15], which we use to estimate $B$ for each virus in each metagenomic study separately. In each study, there are $S$ metagenomic samples taken at various times from $L$ sampling locations. For sample $s \in \{1, ..., S\}$, the data consist of the sampling location $l(s)$, the total number of reads $n_s$, the number of focal viral reads $y_s$, and the public health predictor $x_s$. Because sampling locations vary along several dimensions including sample preparation methods, we use a hierarchical model with a separate term $b_{l(s)} = \log B_{l(s)}$ for each sampling location. The model thus produces a joint estimate of location-specific effects and an overall coefficient for each study and virus.

The Stan model implements the following probability model,

$$y_s \sim Binomial\left(n_s,\ logit^{-1}\left(b_{l(s)} + \theta_s\right)\right) \qquad \text{(A3.3)}$$

$$\theta_s \sim Normal\left(\log x_s, \sigma\right)$$

$$\sigma \sim Gamma(2,\ 1)$$

$$b_{l(s)} \sim Normal(b,\ \tau)$$

$$b \sim Normal(\mu,\ 4)$$

$$\tau \sim Gamma(2,\ 1).$$

where $logit^{-1}$ is the inverse logit (log odds) transformation and μ is a constant determined from the data, as defined below. The first two lines correspond to the model described in Equation A3.2, the next line sets the prior on σ, and the last three lines describe a hierarchical model for the location parameter $b_{l(s)} = \log B_{l(s)}$. Here, $b$ is the overall intercept for the study and virus and τ is the standard deviation of the location-specific terms around this overall value. The prior on $b$ is centered on

$$\mu = \left(\overline{\log x}\right) + \log\left(\overline{y}\right) - \log\left(\overline{n}\right)$$

so that $b = \mu$ roughly represents the naive estimate given by dividing the relative abundance in all of the data by the average log public health predictor estimate across samples. (The priors on $b$, σ, and τ are weakly informative: they constrain the parameters to plausible orders of magnitude, but do not contain any other substantive scientific information. We chose the prior standard deviation of $b$ to be as large as possible while still allowing for efficient posterior sampling (see "Model fitting and checking" below).

The code for the model is publicly available online in GitHub repository associated with the manuscript (see "Data Sharing" in the main text), which includes an annotated version of the Stan code in the model.md 'readme' file.

**Incidence pseudocounts**

Sometimes the reported incidence of a virus is zero for a particular sample. This happens when no new cases were reported in the study region during the period overlapping the sample. To obtain a finite log-incidence for the model, we assigned a pseudocount of 0·1 weekly cases in a given region when incidence for that week & region was zero. Influenza A and B were the only viruses with weeks of zero positive cases, so this adjustment only affected those viruses. To ensure that our choice of pseudocount did not strongly influence the results, we performed a sensitivity analysis varying the pseudocount between 0·01, 0·1, and 1 (Table S11). As expected, the results for SARS-CoV-2 and Norovirus do not change when varying the pseudocount. For Influenza A and B, the results for Spurbeck also do not vary, as Ohio reported cases through all weeks during which we have Ohio wastewater MGS data. The results for Rothman vary slightly, but always well below one order of magnitude. However, for Crits-Christoph, influenza A and B results shift nearly proportional to the value of the pseudocount, representing the fact that close to no influenza A and B were detected in California during the study period of Crits-Christoph. Hence, we dropped Crits-Christoph influenza A and B RA(1%) estimates from Figure 2 and 3.

**Pathogens with zero reads**

Many viruses of interest returned zero mapped reads in some or all of the included studies. Based on our model, such an observation still enables us to estimate weak upper-bound estimates of RA(1%): If two studies sequence at different depths, without detecting a virus with equal prevalence or incidence, the study that sequences more deeply can be expected to have lower RA(1%).

**Model fitting and checking**

We performed model inference using PyStan (24) (v3.10.0). To assess the posterior sampling, we examined plots of the marginal and joint posterior distributions of the parameters. Posterior distributions that have irregular "spikes" at particular values or are strongly multimodal indicate that the Hamiltonian Monte Carlo sampler may not be exploring the parameter space efficiently, likely due to model specification issues. We also compared the prior and posterior distributions to ensure that the priors are weakly informative: they should constrain the posteriors to reasonable values in cases with little data, but have little effect on the posteriors in cases with a lot of data.

These investigations found two issues with our initial choice of priors, which led us to adjust our hyperparameters. First, we found that our initial choice of prior $b \sim Normal(\mu, 2)$ was too narrow, significantly constraining the posterior distribution even for the virus/study combinations with a lot of data. Conversely, we found that $b \sim Normal(\mu, 10)$ was too broad, leading the sampler to mix poorly. We chose the prior $b \sim Normal(\mu, 4)$ as an intermediate value to avoid these issues. Second, we initially used exponential priors for σ and τ, but found that the sampler was colliding with the lower bound at zero. To avoid the boundary, we chose $Gamma(2, 1)$ priors, which have an internal mode at one and vanish at zero.

In addition to visual inspection, we also calculated the R-hat convergence diagnostic for our posterior samples (25). We found that fitting with 8,000 posterior samples on each of four chains gave R-hat < 1.1 for all models (median R-hat < 1.002); we use these settings for all reported results.

We performed posterior predictive checks of the model by generating simulated datasets and comparing the simulated data to the observed data, looking for features not accurately captured by the model. For example, earlier versions of the model did not include terms for sampling locations within studies. Posterior predictive checks revealed that this model failed to predict the low read counts for SARS-CoV-2 in the Point Loma (PL) location of the Rothman dataset, prompting us to add location-specific effects.

#### Relationship between weekly sequencing depth and cumulative incidence at detection

We sought to use our fitted model to understand the sensitivity of a given W-MGS surveillance system to detect a new pathogen with properties similar to the pathogens we created estimates for. We quantify the sensitivity of the system in terms of the cumulative incidence at the time of detection, $C^*$, with lower values of $C^*$ indicating a more sensitive system. We use our model to derive a formula for how $C^*$ depends on the total number of sequencing reads generated per week, $n$. This formula can be inverted to determine how much sequencing is needed to reach a certain $C^*$.

We model detection by supposing that detection of the pathogen occurs when the cumulative number of distinct (non-duplicate) pathogen reads, $Y^*$, crosses a detection threshold $Y^*$. While detection algorithms differ, they all require the presence of sequencing reads originating from the virus of interest. Certain methods—such as mapping to a reference genome to detect a known pathogen—may detect a pathogen from just a few reads, while less targeted methods, such as those involving *de novo* assembly, may require far more. Varying the threshold allows us to represent a range of detection methods.

Imagine a new pathogen with weekly-incidence-to-RA conversion factor $B$ begins spreading in a community with an ongoing W-MGS surveillance system generating $n$ reads per week. Under our statistical model, the expected number of pathogen reads $y(t)$ in week $t$ conditional on the weekly incidence $i(t)$ is

$$E[y(t) \mid i(t)] = n \cdot \frac{B\, i(t)}{1 + B\, i(t)} \approx n\, B\, i(t);$$

the approximation holds for $y(t) \ll 1$, which we assume will be true for a new pathogen that has yet to be detected. Conditional on a given outbreak history (with past weekly incidences of $i(t')$, we can compute the expected cumulative number of pathogen reads seen by week $t$, $Y(t) = \sum_{t' \leq t} y(t')$, by summing over all weeks in the past,

$$E[Y(t) \mid i(t')] = E[\sum_{t' \leq t} y(t') \mid i(t')] \approx \sum_{t' \leq t} n\, B\, i(t') = n\, B\, C(t)\,, \qquad \text{(A3.4)}$$

where $C(t) \equiv \sum_{t' \leq t} i(t')$ is the cumulative incidence at week $t$. In other words, the expected cumulative pathogen reads at a given time is approximately given by the weekly sequencing depth, the weekly-incidence-to-RA factor, and the cumulative incidence, without dependence on the specific incidence history.

In order to use Equation A3.4 to determine the point when detection occurs, we suppose that the cumulative pathogen reads $Y(t)$ at the time of detection is well approximated by the expectation $E[Y(t)]$. This assumption is made for mathematical tractability and warrants future investigation. Setting cumulative pathogen reads equal to the detection threshold $Y^*$ and solving for $C(t)$, we find that the cumulative incidence at the time of detection is

$$C^* \approx \frac{Y^*}{n\, B}\,. \qquad \text{(A3.5)}$$

Intuitively, cumulative incidence at detection is larger (i.e. detection is less sensitive) for higher detection thresholds, smaller amounts of weekly sequencing, and a smaller pathogen weekly-incidence-to-RA factor.

Rearranging Equation A3.5 to solve for $n$ gives an expression for the required weekly sequencing depth required to hit a given detection sensitivity $C^*$,

$$n \approx \frac{Y^*}{B\, C^*}\,. \qquad \text{(A3.6)}$$

Equation A3.6 indicates that the weekly sequencing depth required to hit a given sensitivity grows with a higher detection threshold, lower weekly-incidence-to-RA factor, or higher sensitivity (as measured by $1/C^*$).

*A note about the applicability to scenarios other than the specific studies and pathogens used to estimate the conversion factor B:* The simplicity and generality of Equations A3.5 and A3.6 is a consequence of the fact that the expected cumulative pathogen reads does not depend on the incidence time course, $i(t)$. This fact is a consequence of our statistical model; in particular, its assumption that the expected RA of the pathogen in the sequencing data depends only on its current weekly incidence. In reality, the expected RA may depend on the trajectory of the epidemic in a non-obvious manner, such that a different epidemic time course could lead to a different estimate of B, which implies that estimates of B from the studies in our analysis may not apply to hypothetical new epidemics with different growth trajectories from those in the original estimation studies.

We expect that our estimated values of B can be used so long as either the hypothetical new epidemic follows a similar exponential trajectory as the epidemic used for estimation, or the following two conditions hold for both the estimation and prediction epidemics: 1) shedding during an infection primarily occurs during the several days around when a case is reported and 2) the epidemic doubling time (or halving time, if estimation is done during a time of decline) is on the order of or longer than one week. Under these conditions, weekly incidence is expected to be a good predictor of pathogen RA.

#### Appendix 4: Epidemiological data

##### Epidemiological data

The large differences in viral abundance and composition observed in metagenomic sequencing data could arise from a variety of factors, including differences in sample processing, sewershed hydrology, and the use of DNA vs RNA sequencing. One especially critical factor is the number of people infected with each virus contributing to the sewershed. By accounting for disease incidence and prevalence, we can eliminate this confounder, allowing a more precise evaluation of the performance of W-MGS. Here we discuss further how we arrived at incidence and prevalence estimates for different viruses.

**Virus selection**

To select viruses for which to perform epidemiological estimates, we performed an exploratory literature review choosing viruses based on public-health importance and availability of applicable incidence or prevalence estimates. All included viruses can be found in appendix 2, p 9.

**Data analysis**

For acutely infecting viruses we estimated weekly incidence, while for viruses that cause chronic infections we created estimates of prevalence. All data sources and scripts that process the data to generate incidence and prevalence estimates are available at https://github.com/naobservatory/p2ra-manuscript/tree/main/pathogens. Below, each estimate is explained briefly; Appendix 2, p 9 describes the method of estimate generation for each virus in additional detail.

For incidence viruses, we prioritized the most granular spatio-temporal surveillance data available, starting with daily, county-level testing data. When necessary, we used broader geographic (state or country) and temporal (weekly or monthly) estimates. For SARS-CoV-2 incidence estimates, this resulted in us relying on daily confirmed county-level COVID-19 cases (provided by CDC) (7), smoothed via a centered 7-day moving average, adjusting them upwards by a uniform underreporting factor, also provided by CDC (8). We estimated influenza incidence using state-level, weekly positive testing data provided by CDC (9), which we adjusted by a custom yearly underreporting factor based on the ratio between reported tests (9) and CDC estimates of total symptomatic flu infections (10). Norovirus incidence rates were based on the number of monthly, nationwide outbreaks per year (6), which were transformed into case counts using a 2006 US-wide incidence estimate (5). No incidence estimates were generated for the region and period covered by Brinch (1); this study conducted DNA sequencing, which we expected to not generate sequences for RNA viruses.

For prevalence estimates for these viruses, we prioritized data sources in the following order: (1) recent national-level prevalence estimates; (2) seroprevalence—the share of individuals with antibodies against the virus— or PCR data from national health surveys; (3) prevalence estimates from comparable regions or earlier time periods; and (4) seroprevalence or PCR data from national health surveys in comparable regions or from earlier time periods.

For the United States, publicly available prevalence estimates were available for human immunodeficiency virus (HIV) (11), hepatitis C virus (HCV) (13), and herpes simplex virus 1 (17). When prevalence estimates weren't available, we used public seroprevalence data instead. This was the case for cytomegalovirus (CMV) (19), and Epstein-Barr-virus (EBV) (15). PCR testing data was available for human papilloma virus (HPV) (21) in 15-59 year olds. Both seroprevalence and PCR data was sourced from publicly available datasets collected in the National Health and Nutrition Examination Survey

(NHANES), a biannual health survey of a US population sample that includes testing for common chronic infections (26). For viruses of lower public-health concern, estimates were not generally available from either CDC or NHANES sources discussed above.

For Denmark, publicly-available prevalence estimates were available for HCV (13) and HIV (12). For EBV (16) and HPV (22), we used published seroprevalence and qPCR positivity estimates, respectively, as proxies for overall prevalence. When in-country estimates weren't available, we resorted to the best data source from a European country or region with similar demographics (Germany for HSV-1 (18), the Netherlands for CMV (20)).

#### Appendix 5: Metagenomic data analysis

FASTQ files for each included study were obtained from the Sequencing Read Archive (27) and analyzed with a custom computational pipeline (see "Data Sharing") as follows:

1. Raw reads were screened for adapter contamination with Cutadapt (28), Trimmomatic (29), and FASTP (30) Additionally, FASTP was used to trim low-quality and low-complexity sequences.
2. Cleaned reads underwent deduplication with Clumpify (31).
3. Deduplicated reads were ribodepleted with BBDuk (32), using SILVA SSU and LSU sequence databases, version 138.1 (33).
4. Ribodepleted reads were then separately analyzed in a taxonomic profiling and a human-infecting virus identification pipeline. In the taxonomic pipeline, paired-end reads were merged with BBMerge (34), with reads that failed to merge being concatenated with an intervening "N" base. Sequences were then passed to Kraken2 for taxonomic assignment, using the Standard database (35) (2022-12-01 build), then summarized with Bracken (36).
5. The human-infecting virus pipeline included the following steps:
    a. Beforehand, a database of human-infecting viral genomes was generated by obtaining all human-infecting virus taxonomy identifiers from Virus-Host DB (37); expanding this list to include all descendant identifiers; downloading all viral genomes corresponding to these identifiers from Genbank (38); and filtering the resulting database to remove transgenic and contaminated sequences.
    b. Ribodepleted reads were aligned against this database with Bowtie2 (39) to identify putative human-infecting virus reads. Each read is assigned an NCBI taxonomy ID (taxid) corresponding to the best alignment found by Bowtie2.
    c. Putative human-infecting virus reads were filtered by aligning them to reference genomes that include human, cow, pig, mouse and *E. coli*, as well as various genetic engineering vectors. Alignment was performed by Bowtie2 and BBMap (40) in series.
    d. After filtering, read pairs were merged with BBMerge and taxonomically assigned with Kraken2 as above. Each read was either (1) assigned to a human-infecting virus taxon with Kraken, (2) assigned to a non-HV taxon with Kraken, or (3) not assigned to any taxon. All reads in category (2) were filtered out.
    e. Reads are assigned a HV status if (i) they are given an HV assignment by both Bowtie2 and Kraken2; or if (ii) a read is unassigned by Kraken but aligns to an HV taxon with Bowtie2 with an length-normalized alignment score above a specific user-defined threshold of 20 (i.e. alignmentScore/ln(readLength) >= 20).
    f. The number of reads assigned to each human-infecting virus taxon are calculated by summing all Bowtie2 assignments to that taxid and its taxonomic descendants, according to the NCBI taxonomy hierarchy. These read counts were then used to calculate RA(1%) estimates as described in Appendix 3. The taxids used to generate such estimates are documented in Table S5.

#### Appendix 6: Cost modeling

##### Overview

We consider a surveillance approach $j$ with one wastewater sample per week, giving 52 samples a year. The total per-sample cost $c_j$ will be made up of the per-sample **processing cost** $c_j^{proc}$ (including, for instance, library prep, rRNA depletion, or probe-hybridization enrichment) and the per-sample **sequencing cost** $c_j^{seq}$:

$$c_j = c_j^{proc} + c_j^{seq} \quad \text{(A6.1)}$$

We then calculate the annual cost $C_j$

$$C_j = c_j \times 52 \text{ weeks} \quad \text{(A6.2)}$$

Each surveillance approach $j$ is defined by a particular combination of processing steps, detection threshold, and incidence-to-RA factor. Per-sample processing costs $c_j^{proc}$, sequencing costs $c_j^{seq}$, total costs $c_j$, and annual total costs $C_j$ are given in Table S10.

##### Per-sample processing costs

The per-sample processing cost $c_j^{proc}$ consists of the cost of library preparation $L$, as well as the costs of rRNA depletion $R$ and panel-enrichment $P$ for those surveillance approaches that include them. $L$, $R$, and $P$ are given like so:

$$R_j = \begin{cases} R & \text{if approach } j \text{ uses rRNA depletion,} \\ 0 & \text{otherwise,} \end{cases} \qquad P_j = \begin{cases} P & \text{if approach } j \text{ uses panel enrichment,} \\ 0 & \text{otherwise.} \end{cases}$$

For a surveillance approach $j$, this gives,

$$c_j^{proc} = L + R_j + P_j \quad \text{(A6.3)}$$

rRNA depletion is only used in Crits-Christoph, and panel-enrichment only in panel-enriched samples from Crits-Christoph and Rothman. Processing costs are applied irrespective of sequencing depth.

Library preparation costs $L$ were sourced from public pricing data provided by the Harvard Bauer Facility, using the "Harvard Account Code" costs which represent the cost to university affiliates (41). The per-sample library preparation costs amount to $L$=\$216. Ribodepletion cost $R$ is based on the cost of the Illumina Ribo-Zero Plus rRNA Depletion kit(42), which at the time of writing is \$910.00 for 16 samples, giving a per-sample cost of $R$=\$57. Similarly, panel enrichment cost $P$ is based on the Illumina Respiratory Virus Enrichment Kit Set (43), with costs of \$10,368.00 for 32 reactions, resulting in a per-reaction cost of $P$=\$324. Only costs specific to W-MGS (as opposed to targeted methods like qPCR) were considered.

The library preparation costs $L$ are fully-loaded service costs which include labor. Conversely, the ribodepletion costs $R$ and panel-enrichment costs $P$ are materials-only costs which exclude labor. The total sample processing cost $c_j^{proc}$ will thus underestimate the labor costs involved in sample processing. However, we expect the impact of this underestimation on our final cost estimates to be small, for two reasons. First, the labor involved in ribodepletion and panel-enrichment is small compared to that involved in library preparation, which is a complex multi-stage process. Second, the amount of additional labor required to add these steps is consistent across sequencing platforms, as all involve the same methods and materials being applied in the same way.

#### Per-sample sequencing costs

Per-sample sequencing costs $c_j^{seq}$ were derived as follows. First, the required weekly sequencing depth $n_j$ was calculated based on a study- and pathogen-specific weekly-incidence-to-RA factor $B_j$, a detection threshold $Y_j$, and a target detection sensitivity $C_j$, as previously in Equation 3.6 like so:

$$n_j \approx \frac{Y_j}{B_j C_j} \qquad \text{(A6.4)}$$

Subsequently, the sequencing costs $c_j^{seq}$ for reaching the required sequencing depth $n_j$ were calculated for four different sequencing approaches $k$, each with a different per-unit cost κ, and estimated read output Ω:

- **MiSeq Micro v2 300 cycle flow cell**
  - $\kappa = \$788$
  - $\Omega = 2\times10^6$ read pairs
- **NextSeq 1000 P1 300 cycle flow cell**
  - $\kappa = \$1{,}397$
  - $\Omega = 4.5\times10^7$ read pairs
- **NovaSeq X Plus 25B 300 cycle flow cell (per-lane)**
  - $\kappa = \$2{,}494$
  - $\Omega = 1.4\times10^9$ read pairs
- **NovaSeq X Plus 25B 300 cycle flow cell (full flow cell)**
  - $\kappa = \$18{,}902$
  - $\Omega = 2.34\times10^{10}$ read pairs

Per-unit costs κ and read output Ω were obtained from the Harvard Bauer Facility, using the "Harvard Account Code" costs (41).

For a given surveillance approach $j$, the sequencing cost associated with a given sequencing approach $k$ was determined based on the minimum integer number of units (flow cells or lanes) required to meet or exceed the target sequencing depth $n_j$:

$$c_{j,k}^{\mathrm{seq}} = \kappa_k \times \left\lceil \frac{n_j}{\Omega_k} \right\rceil \qquad \text{(A6.5)}$$

Finally, the overall sequencing cost $c_j^{seq}$ was calculated as the minimum over all sequencing approaches k:

$$c_j^{\mathrm{seq}} = \min_k \left\{ c_{j,k}^{\mathrm{seq}} \right\} \qquad \text{(A6.6)}$$

https://journals.plos.org/plosone/article?id=10.1371/journal.pone.0238203

36. Lu J, Breitwieser FP, Thielen P, Salzberg SL. Bracken: estimating species abundance in metagenomics data. PeerJ Comput Sci. 2017 Jan 2;3(e104):e104.
